# Categorising Coercion in Assisted Dying. An AI-Assisted Analysis of the House of Commons Debate on the 2025 Terminally Ill Adults Bill in England and Wales

**DOI:** 10.1101/2025.09.12.25335636

**Authors:** Natasia Hamarat, Juan González-Hijón, Jacques Wels

## Abstract

**Background:** The debate on voluntary assisted dying (VAD) in England and Wales has long centred on safeguarding autonomy and preventing coercion.

**Methods:** This study analyses parliamentary debates (2024–2025) on the Terminally Ill Adults (End of Life) Bill to explore how Members of Parliament (MPs) framed coercion risks in assisted dying legislation. Using AI-assisted text analysis of Hansard transcripts, we identify seven coercion types: economic (financial pressure), ethnic (healthcare inequalities), familial (family influence), medical (clinician bias), mental (psychiatric conditions), poor care (inadequate palliative services), and self-coercion (internalised pressure).

**Results:** These concerns were transpartisan, with no party disproportionately emphasising any single coercion type. Poor care and mental coercion were most frequently cited, reflecting concerns over UK healthcare underfunding and psychological vulnerability. Network analysis revealed strong co-occurrence between medical and mental coercion, as well as familial and economic pressures, suggesting MPs viewed these as interconnected risks. However, debates largely focused on individual-level safeguards (e.g., capacity assessments) rather than systemic monitoring of structural inequalities.

**Conclusions:** We argue that effective VAD regulation requires a dual approach: individual protections alongside population-level oversight to address disparities in access and prevent coercion across socioeconomic and ethnic groups. This framework could inform future legislation in the UK and other jurisdictions considering assisted dying reforms.

## Background

As of 2025, England, Wales and Scotland had no legislation on assisted dying. By contrast, neighbouring states such as the Netherlands [1], Belgium [2] (both in 2002), Switzerland (1941) [3] and, more recently, Spain (2021) and Portugal (2023), have introduced various forms of voluntary assisted dying (VAD), including euthanasia and assisted suicide. In England and Wales, assisted dying is prohibited by the 1961 Suicide Act, later amended by the 2009 Coroners and Justice Act [4]. Many Bills have been introduced in recent years to change the law, including those in 2004 [5] and 2006 [6], in 2014, in 2015, and in 2021 [4], but all have failed. Due to devolution, England and Wales, Scotland and Northern Ireland may pass different legislation on the matter [7]. While there is evidence of public support for the implementation of such legislation [8], the Scottish Assisted Dying for Terminally Ill Adults Bill was introduced in March 2025 [8] and, in October 2024, the Terminally Ill Adults (End of Life) Bill was introduced to Parliament for England and Wales.

In June 2025, the Bill passed its third reading and moved to the House of Lords. At this stage, the *Bill* permits mentally competent adults (18+) with a terminal illness (≤6 months prognosis) to request assisted dying, excluding cases based solely on disability or mental illness. Safeguards include assessments by two doctors and a multidisciplinary panel (lawyer, psychiatrist, social worker) to confirm the decision is voluntary, informed, and free from coercion, alongside mandatory reflection periods – 7 days between medical assessments and 14 days (or 48 hours if death is imminent) after panel approval.

One key feature of these Bills is that they are *transpartisan*: no major party took an official position on the matter, and MPs were not required, as is the case with other regulations, to follow their party’s line. This has not always been the case and might explain the more favourable outcome observed in 2025. In 2015, there were elements of party influence in the debate with, for instance, conservative MPs following the line of the party leader [9]. Party was not a main source of influence but it did matter in some indirect ways.

The principal arguments for or against previous Bills were consistently articulated around several key notions [10]: palliative care, autonomy, choice and control, legal and social effects [10]. The potential coercive effects of VAD regulations have long been at the heart of the debate. In 2021, for example, the term ‘pressure’ was discussed in 23% of all parliamentary speeches [10], referring notably to forms of pressure exerted by family members or broader societal expectations. The word ‘coercion’ was mentioned in 8% of speeches. Between 2014 and 2022, most concerns regarding VAD legislation focused on the potential pressure on individuals [4].

At the heart of both the Scottish and the English-Wales Bills lies the notion of coercion [11]. The question of autonomy/coercion in VAD has been put into question from both an empirical and an ethical point of view [12]. From an ethical standpoint, it has been argued that autonomy in assisted dying faces the paradox that the choice to request it depends upon the fact that there would be no alternative, and assisted dying is seen as an instrument of force for vulnerable people facing difficult end of life choice [13]. Within this view, improving end-of-life care access and provisions [14] and avoiding coercion rather than extending the spectrum of VAD regulations appear as key policy issues [15]. Such positions are debated in the scientific literature and whether people are not competent in choosing euthanasia because of pain, anxiety or desperation and whether these legislations would lead to social pressure on patients have not been evidenced by empirical studies [16]. Rather, empirical studies show that the primary reason for assisted dying is the unbearable suffering experienced by the patients, which, in this perspective, should help policymakers understand the need for such regulations [17]. Nevertheless, most of the debate when implementing some kind of VAD regulations focuses on whether the choice is made freely and voluntarily and the need to carry out capacity assessment and address potential undue influence [18, 19] – coercion, not autonomy, emerges as the central concern.

## Methods

### Aim

Using House of Commons transcripts (Hansard) of the recent 2024-25 Bill in England and Wales, this study has three objectives. (1) First, to examine the narratives around coercion used by MPs to frame the factors that could compromise autonomy, as perceived by the people’s representatives, and to develop a typology of these potential coercive factors. (2) Second, to explore the transpartisan nature of the debate by quantifying how the different types of coercion are distributed across parties. (3) Third, to map a network of coercion types to understand how these forms of coercion are articulated together in MPs’ narratives. By doing so, the study pursues a dual objective: (a) to provide an overview of the factors that could affect patients’ autonomy, as perceived by policymakers, in order to clarify which factors should be monitored in the future; and (b) to offer a framework to inform potential debates in other nations or states considering VAD regulations.

### Data Collection and Pre-processing

This study analysed the official Hansard transcripts of the House of Commons debates on the Terminally Ill Adults (End of Life) Bill 2025, as published in the parliamentary record. The transcripts were pre-processed to isolate individual parliamentary interventions by splitting the text at each speaker transition marker (“#”). Speech segments shorter than 30 characters were excluded to retain only substantive contributions. The three readings (16 October 2024, 29 November 2024, and 20 June 2025) yielded 118 valid speech segments. The committee stage, comprising 29 sittings held between January and March 2025, resulted in 4,657 valid interventions, grouped by speaker. A summary of the sessions is provided in **Table 1**; full details on all sittings and readings are available in <u>Supplementary File S.1</u>.

**Table 1.** Timeline summary

| Stage | Date | Key Outcome | Vote Margin |
| --- | --- | --- | --- |
| First Reading | 16 Oct 2024 | Bill presented to Commons | N/A |
| Second Reading | 29 Nov 2024 | Approved for committee scrutiny | 330–275 |
| Committee Stage | Jan–Mar 2025 | 500+ amendments debated over 29 sittings | N/A |
| Report Stage | May–Jun 2025 | Safeguard amendments adopted | Mixed votes |
| Third Reading | 20 Jun 2025 | Final Commons approval | 314–291 |
| House of Lords | Late 2025–2026 | Ongoing scrutiny | Pending |

### AI-Assisted Text Classification

Each speech segment was processed using DeepSeek-V3’s natural language API (DeepSeek-chat model). Application Programming Interface (API) refers to a defined set of protocols that allows different software systems to communicate, enabling automated submission of text data to the AI model and retrieval of structured outputs for analysis.

The first application of the model aimed to develop a typology of coercion arguments based on the Bill’s reading sessions. This was achieved through sequential classification of segments for: (1) speaker identity; (2) party affiliation; (3) position on the bill (for or against); (4) presence of coercion-related arguments (including synonymous terms); and (5) exact phrasing of coercion arguments. Building on these outputs, the authors carried out an inductive thematic analysis, completed by a close reading of the debates. Through interpretative coding, they identified ideal types recurring patterns which were identified and progressively shaped into a seven-category types of coercion arguments [20]. A second application of the API used this typology to systematically identify and extract mentions of coercion type by speaker across the debates in both the reading and siting sessions. In the reading stage, where speakers typically intervened once per session, coercion type identification was directly linked to each speech. During the committee stage, where speakers could intervene multiple times during a sitting, coercion mentions were aggregated by speaker within each sitting. The API was configured with temperature = 0 to ensure deterministic outputs [21], with responses limited to 200 tokens to maintain focus. A consistent prompt engineering strategy enforced semicolon-delimited output formatting, which was cleaned and harmonised for analysis.

The classification underwent a two-stage validation process. First, technical validation involved logging API errors and assigning NA values to incomplete responses. Second, manual review of a random 20% sample of the data assessed the accuracy of classifications, with particular focus on the identification of coercion arguments and types.

### Analysis

Descriptive analyses were conducted separately for the reading stages and committee stage to explore patterns in the use of coercion arguments.

First, coercion type variables were converted to numeric indicators (presence = 1, absence = 0). Data were filtered to focus on major political parties (Conservative, Labour, Liberal Democrat, Democratic Unionist Party) and valid speech records.

For the reading stages and committee stage, frequency distributions of coercion types were calculated by parliamentary stage (reading or committee sitting). These were visualised as stacked bar plots displaying the proportional contribution of each coercion type within a given stage.

To examine patterns by political affiliation, we calculated the within-party distribution of coercion types (i.e. the relative focus each party placed on different coercion types given its total mentions of coercion) and displayed results using stacked bar charts. All visualisations were formatted for clarity, with percentage labels displayed where values exceeded 10 percent.

Finally, to explore how coercion types co-occurred within speeches, pairwise correlations between coercion type indicators were computed (using pairwise complete observations). Correlations greater than 0.15 were retained to generate an undirected co-occurrence network. This network was visualised using force-directed layout graphs, where edge thickness and opacity reflected correlation strength and node labels indicated coercion type.

All analyses were conducted in R (version 4.3.2), using the *httr* package for API interactions and *tidyverse*, *igraph* and *ggraph* for data management and analysis.

## Results

### Seven types of coercion

Preliminary analyses of the three reading sessions distinguish seven potential sources of coercion in the MPs interventions: economic, ethnic, familial, medical, mental, poor care and self-coercion. Table 2 provides examples of the different coercion types mentioned throughout the reading sessions.

**Table 2.** Coercion sources and Hansards relevant extracts.

| Coercion type | Hansards relevant extracts |
| --- | --- |
| Economic | “This is an equity issue: if a person is financially poor, terminally ill and nearing the end of their life, they do not have the freedom to choose assisted dying. That does not mean, of course, that they would, but this legislation will allow an equitable choice for this defined group of people.” **Dr Beccy Cooper, Labour MP, Third Reading**<br>“I ask colleagues in favour of passing the Bill to consider the following scenario. An older relative knows that assisted death is now possible and that their family is struggling to get by, in difficult economic circumstances. They have a health condition, with a prognosis of five months to live, even though studies show that most such prognoses are wrong about 50% of the time. What will stop our parents or grandparents from deciding to seek assisted dying purely to “do the right thing” by their loved ones?” **David Smith, Labour MP, Third Reading** |
| Ethnic | “We should specifically consider the impact on ethnic communities: we know the prism of racist assumptions through which healthcare has too often been administered —the huge inequalities in maternal health and mental health, to name just two examples. There is nothing in this Bill to protect the vulnerable and those whose experience of life and death has already been biased.” **Dame Chi Onwurah, Labour MP, Third Reading**<br>“All the data shows us that people from minority ethnic communities are less likely to access the healthcare services they deserve, in particular palliative care. There is a deep distrust of health services, and those of us who were in this place during the covid pandemic saw that played out in real time in hospitals and care settings up and down the country, with far more people from minority ethnic communities losing their lives and far more healthcare professionals from ethnic minorities not protected in the way that they should have been.” **Munira Wilson, Liberal Democrat MP, second reading** |
| Familial | “I was contributing an observation from somebody who has been deeply involved in palliative care practice, who reports that it is far more frequent that the dying person wishes to die, while it is their family who are pressuring them and encouraging them to stay alive as long as possible. The fears about coercion appear to be worry* about something that is not actually the case in these cases of dying people.” **Ellie Chowns, Green MP, Second reading** (*note: typo comes from the Hansard)<br>“There has been a lot of talk about there being no evidence of coercion, but within the family, the most powerful coercion is silence: it is the failure to answer when a question is put. If police cannot spot coercion in domestic violence, how can they be expected to spot coercion in assisted dying?” **Diane Abbott, Labour MP, Third Reading** |
| Medical | “I do not believe that we are stepping on to a slippery slope or unpicking the very purpose of the NHS, as some have suggested. We are here simply to give those who already face terrible decisions—doctors, patients and their families—a real choice of how to face those decisions, and protection in law for choices that are already being made today.” **Luke Taylor, Liberal** |
|  | **Democrat MP, Third Reading**<br>“Imagine that you have a brother who has struggled with an eating disorder ever since he entered secondary school. He was sexually abused by a family friend and never received any real support for what he went through, and court backlogs mean that the criminal case is still ongoing. He spends longer and longer watching social media influencers paid by assisted dying companies to advocate for what they call a peaceful end to life. He begins to starve, and doctors withdraw treatment because they claim nothing more can be done. You get a call only a few months after his 18th birthday to tell you that your brother has opted for an assisted death.” **Jess Asato, Labour MP, Third Reading** |
| Mental | “People with anorexia stop eating and drinking because they have a psychiatric illness. Those are two categorically different issues. I must make it absolutely clear that even though amendment 14 has passed today, it does not address concerns about anorexia or close that loophole.” **Naz Shah, Labour MP, Third Reading**<br>“What assurances does the Bill give to the families of people with a disability, or those with mental health issues and those who are anorexic? I do not see any. Does the hon. Lady see any assurances for those who want to end their lives but suffer from those ailments?” **Jim Shannon, DUP, Second Reading** |
| Poor care | “[...] our palliative care sector provides high-quality, compassionate and dignified care at its best, access to palliative care across the country is, sadly, not at a uniform level for all people who need it at the end of life. Sadly, in terms of funding and delivery, the majority of palliative care is left to the charitable sector.” **Dr Neil Hudson, Conservative MP, Third Reading**<br>But what choice does the Bill hold for someone who, all their life, has lacked agency, particularly in a family context, which may be particularly the case in certain cultures and communities? And what choice does the Bill offer to those who lack access to good palliative care? **Diane Abbott, Labour MP, Third Reading** |
| Self | “Why do we want to restrict advertising about assisted dying? It is not just because such adverts could appear crass or insensitive, or because we worry that private companies could profiteer from death, but because advertisers know that they influence choices. The issue of choice, whether it is informed choice, skewed choice, self-coercion or coercive control, as has already been mentioned, is, in many ways, at the heart of the Bill and whether its safeguards are sufficient.” **Paul Waugh, Lab/Co-op MP, Second Reading**<br>“The hon. Member will be aware that the Bill creates a criminal offence that would punish those who would coerce a relative in such a way. [Hon. Members: “Self-coerce.”] There are folks who talk about the concept of self-coercion, but others would frame such a decision as a choice. Self-coercion is a choice.” **Josh Babarinde, Liberal Democrat MP, Third Reading** |

*Economic coercion* refers to a situation where a person’s financial hardship or poverty limits their freedom to make a genuine, autonomous choice about assisted dying. The fear is that terminally ill people who are poor may feel pressured—whether overtly or subtly—to choose assisted death to avoid being a financial burden on their families or the state.

*Ethnic coercion* arises from the systemic inequalities, biases, and racism that ethnic minority communities may face within the healthcare system. Because of historic and ongoing discrimination, people from these communities might distrust healthcare services or fear that their care will be inadequate or biased, making them more vulnerable to feeling they have no alternative but to choose assisted dying.

*Familial coercion* refers to pressure that comes from family members. This could take the form of direct encouragement to choose assisted dying, subtle signals that death would ease the family’s burden, or even the powerful silence of unspoken expectations. It acknowledges the complex emotional dynamics that can exist within families at the end of life.

*Medical coercion* occurs when healthcare professionals—intentionally or not—exert influence over a patient’s decision about assisted dying. This could involve presenting assisted death as the most practical or compassionate option, particularly when resources are limited, or when the patient feels abandoned or dismissed by the medical system.

*Mental coercion* refers to the impact of a person’s psychiatric or psychological condition on their decision-making capacity. For example, individuals suffering from severe mental health conditions such as depression, anorexia, or trauma may seek assisted dying not because of an autonomous wish to die, but as a result of their illness influencing their judgement.

*Poor care coercion* happens when inadequate access to quality palliative care or end-of-life support leads a person to see assisted dying as their only dignified option. The lack of good care, rather than the desire for death itself, may drive their decision.

*Self-coercion* refers to the internalisation of external pressures or societal messages that lead a person to convince themselves that assisted dying is the right or necessary choice. This could stem from feeling like a burden, absorbing cultural or commercial messages, or experiencing internal conflict, rather than making a free and informed decision.

### Coercion type focus by reading and sitting sessions

During the First Reading on 16-Oct-24, 31 speeches were analysed, with 12 in favour of the bill and 18 against, while coercion was mentioned 21 times. In the Second Reading on 29-Nov-24, 42 speeches were analysed, with 21 in favour and 20 against, and coercion mentioned 14 times. The Third Reading on 20-Jun-25 had 43 speeches analysed, with 19 in favour and 24 against, and coercion mentioned 28 times. Across all readings, a total of 116 speeches were analysed, with 52 in favour of the bill, 62 against, and coercion mentioned 63 times (see <u>supplementary file S.2.1 and S.2.2</u>).

Over the three readings, coercion due to poor care or mental issues (including psychological or psychiatric) were mentioned by respectively 30 and 26 speakers. Family coercion was mentioned 21 times. Economic coercion and coercion by medical staff were both mentioned by 18 speakers. Self-coercion and coercion because of ethnicity were mentioned respectively by 17 and 11 speaker (see <u>supplementary file S.2.3</u>). **Figure 1** shows the distribution (in percentage) of these coercion types by reading stage and party affiliation (focusing on parties that made more than 5 interventions). On the left hand size where each debate stage totals 100 percent, it can be observed that coercion types were consistent across the different stages with, for instance, poor care mention by 22, 18 and 23 percent of the speakers in first, second and third readings and coercion due to medical practitioners respectively mentioned 12, 11 and 14 percent of the speakers. On the right hand size, the same finding appears for party affiliation: the relative focus on different coercion types did not appear to be affected by party affiliation.

**Figure 1.**
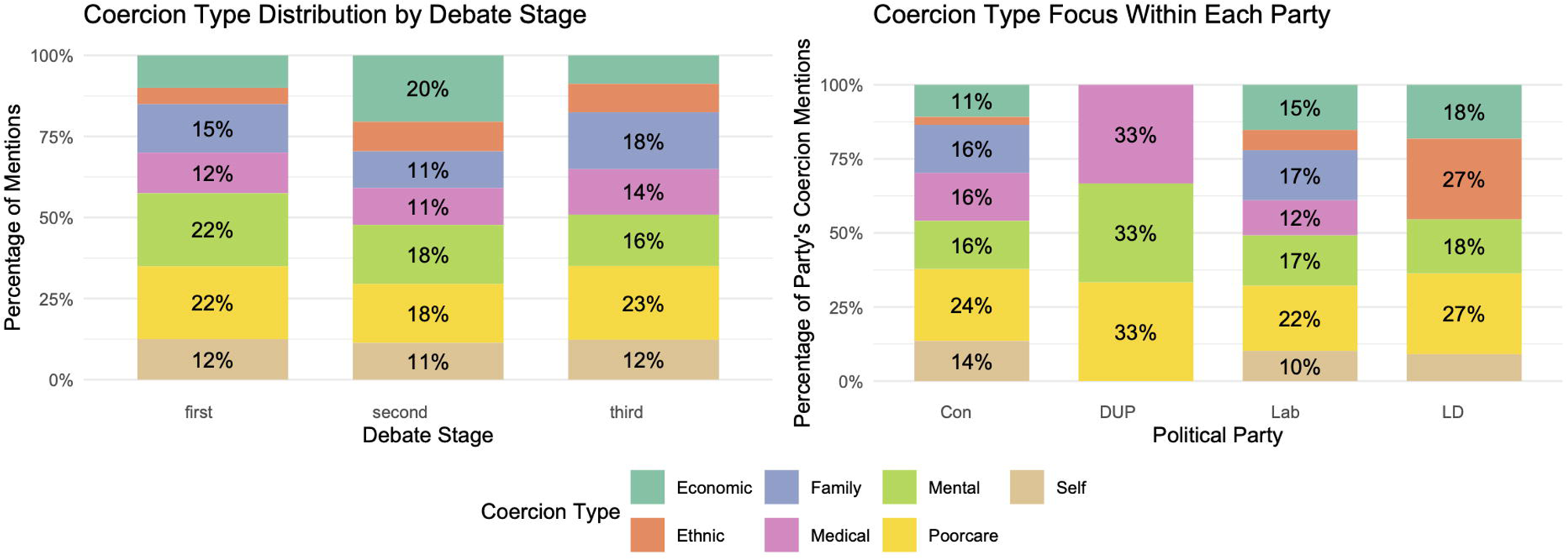
Coercion type focus by reading session and party

In the thirty sessions of the sitting stage (including the introductory sitting), a total of 4,657 interventions were recorded, as shown in <u>Supplementary File S.2</u>. Mental coercion was mentioned by 144 speakers (i.e., total count of speakers including repeated individuals), and family coercion was mentioned by 97. Poor care, economic and self-coercion were mentioned by 63, 62 and 53 speakers respectively. Ethnic coercion was mentioned by 33 repeated speakers. **Figure 2** replicates the descriptive analysis for sitting stage. The right-hand panel shows that all coercion types were mentioned during the sitting stage, although some were discussed more frequently than others. *Mental coercion* was mentioned most extensively, appearing in 29 of the 30 sessions and accounting for between 7 percent (1^st^ sitting) and 73 percent (19th sitting) of the coercion-related arguments. *Poor care coercion* was also discussed during the sittings, although to a lesser extent overall; notably, the 26th sitting focused almost entirely on the implications of poor care. *Self-coercion* was mentioned throughout the sitting stage as well, with the exception of four sessions where it did not arise.

**Figure 2.**
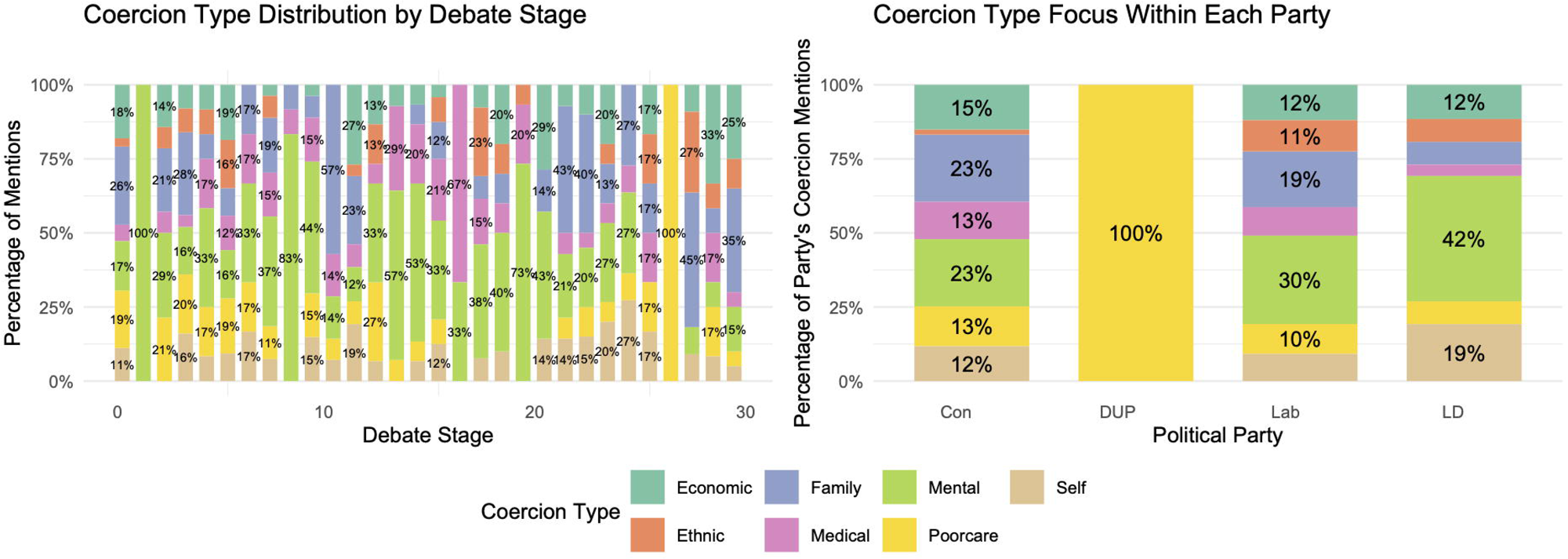
Coercion type focus by sitting stage and party

Although the Committee Stage, conducted by a 23-member Public Bill Committee, was formally apolitical in composition (reflecting proportional party representation), the distribution of coercion types observed in the sittings mirrored that of the reading sessions: all types were nearly equally distributed across parties. Notably, mental coercion was more frequently cited in both sittings and readings, whereas poor care coercion appeared less often in the sittings compared to the readings. Note that only one member of the DUP was involved in the sitting which led to only one coercion type observed in figure 2.

### Coercion type co-occurrence

Figure 3 shows the co-occurrence networks for the coercion type indicating how often correlation types were mentioned together by the different speakers. The matrices of correlations used in figure 3 are shown in <u>supplementary file S.4</u>.

**Figure 3.**
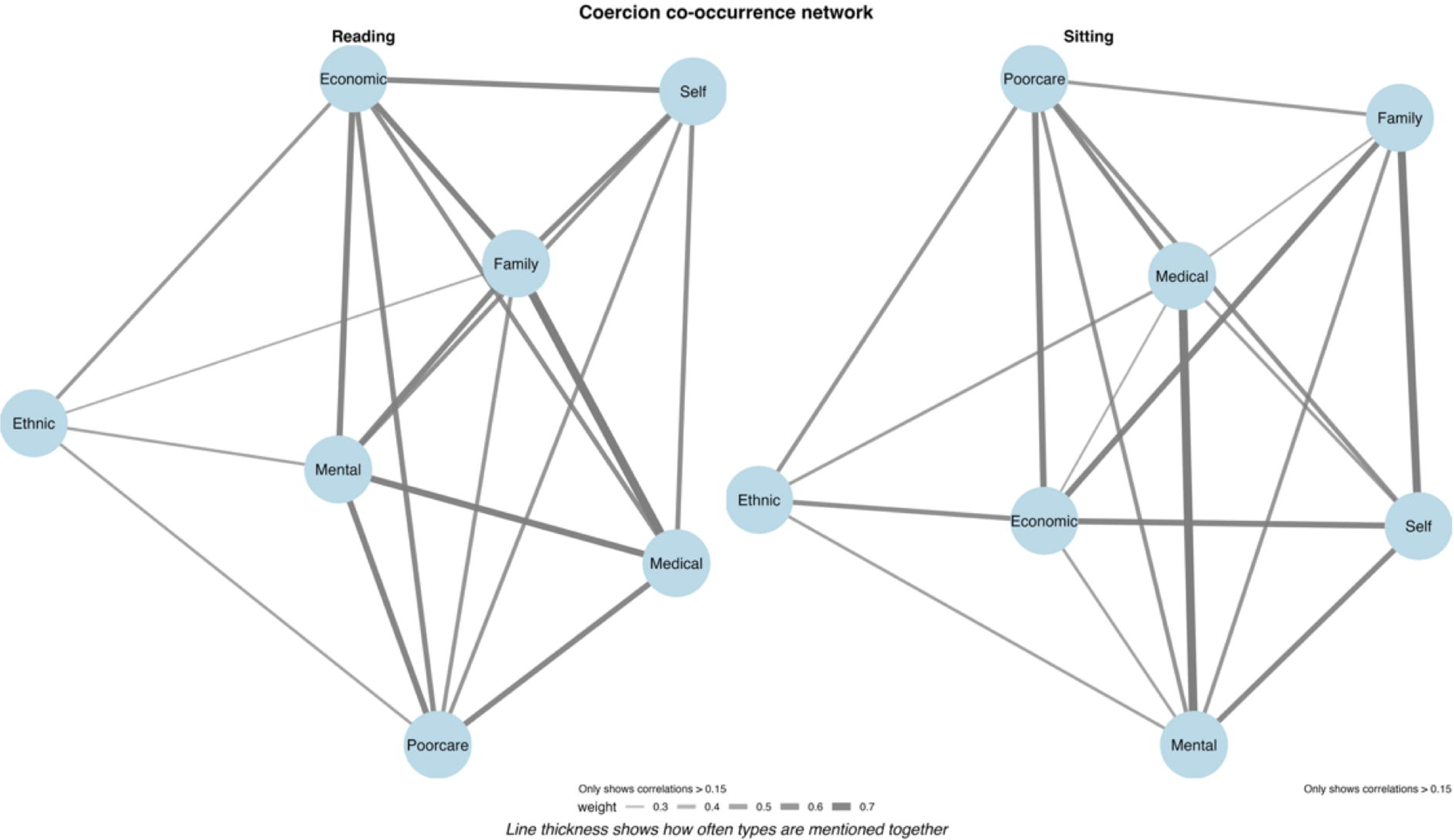
Coercion type co-occurrence network in the HC reading and sitting sessions

The figure shows differences between how coercion types are connected in brief statements (Reading) versus extended debates (Sitting). In the Readings, speakers tend to combine multiple coercion types together, showing particularly strong connections between family and medical coercion (r = 0.76), as well as between medical and mental coercion (r = 0.60). Economic coercion also shows moderate to strong links with several other types, including family (r = 0.57), mental (r = 0.57), and self (r = 0.52). These patterns suggest that in short statements, speakers frequently mention these coercion types together as part of broader narratives.

The Sitting debates show much weaker connections overall. The previously strong family-medical link drops dramatically (r = 0.17), and no correlations exceed 0.49 (medical-mental r = 0.49). Economic coercion maintains only modest associations with other types (family r = 0.36, poor care r = 0.36, self r = 0.35). Self-coercion the exception, maintaining similar moderate connections in both contexts (family r = 0.50 in Reading vs. 0.45 in Sitting; economic r = 0.52 vs. 0.35).

These findings demonstrate how discussion format shapes the presentation of coercion. Brief statements tend to combine multiple coercion types, creating stronger statistical connections. In longer debates, speakers examine each coercion type more separately, resulting in weaker correlations. The particularly large drop in the family-medical connection (from 0.76 to 0.17) highlights this difference most clearly. Only self-coercion maintains consistent connections across both contexts, suggesting it may serve a unique role in discussions of coercion.

## Discussion

Access to VAD is becoming more widely available around the world [22, 23]. The UK Parliament was debating the implementation of an assisted dying regulation in England and Wales in 2024-25 with the legislation likely to be implemented in the coming years. In such a context, this study analysed the content of the House of Commons debate and more specifically focuses on the way the notion of coercion was framed by parliamentary members.

In this study, we identified seven types of coercion mentioned by MPs. Coercion is perceived to stem from economic circumstances, family, mental health (psychological or psychiatric), ethnic background, relationships with medical practitioners, poor care, and self-coercion. The study highlights the transpartisan nature of the coercion narrative: all types of coercion are mentioned by MPs across all parties with poor care and mental coercion appearing most frequently. These reflected concrete UK policy concerns, particularly regarding healthcare system vulnerabilities. The consistency across readings shows these concerns remained central throughout legislative scrutiny. The distribution of coercion types showed notable consistency across party lines. Conservative, Labour and Liberal Democrat speakers emphasised similar proportions of each coercion type. This suggests that decisions regarding the implementation of assisted dying in England and Wales are shaped more by individual beliefs or informed by external bodies than by party affiliation. In the late stage of the debate, medical bodies such as the Royal College of Psychiatrists (RCP) – although the role of psychiatrists as social gatekeepers has been criticised [24] – have underlined the role social relationships and people’s need in shaping VAD decision [25, 26].

Network analyses revealed stable relationships between coercion types throughout the debates. Medical and mental coercion showed particularly strong co-occurrence, while family coercion correlated moderately with both economic and medical coercion. These patterns persisted across readings despite evolving amendments, suggesting MPs consistently viewed certain coercion types as interrelated. Yet, these forms of coercion refer to different levels of the social reality that deserve specific forms of attention and monitoring. Family, medical, mental and self-coercion refer to a *relational* level where VAD is seen as the product of the relationships with others [27, 28]. In such a perspective, the relationships family and care practitioners would shape the decision to die [27]. Poor care refers to contextual factors: these are seen as determinants patients have no grasp on and that would affect the decision to request VAD in a context of underfunding of palliative care [14, 29]. Finally, the impact of the ethnic background or economic factors refers to the potential structural determinants of VAD. Whilst no study has demonstrated that some specific populations would be more prompt to access VAD in other countries [23, 30], these factors were constantly mentioned in the debate.

## Conclusion

Different levels of coercion should point out towards different forms of control of any assisted dying regulation. The current debate on assisted dying and euthanasia regulations in the UK and France largely focuses on individual-level safeguards [4]. Both proponents of stricter controls and advocates for broader access concentrate on how requests are assessed, by whom, and whether juridical or psychiatric oversight is required. While these discussions on case evaluation are important, the repeated mentions of contextual and structural factors indicate that VAD regulations should also be approached as a public health policy and it is surprising that population-level monitoring is largely absent from the debate. The risk is to evaluate individual requests whilst not addressing broader population-level trends and inequalities. This echoes the insights of Geoffrey Rose in his paper, *“Sick Individuals and Sick Populations”* [31], where he argues that health interventions must operate on two levels. While individual-level approaches respond to the needs of specific patients, population-level strategies are necessary to understand and mitigate the wider social, economic, and structural factors that shape health outcomes. Although poor care and economic and ethnic inequalities have been highlighted by many MPs throughout the legislative process, these issues have received comparatively less attention in terms of safeguards and data monitoring. The argument works in both directions: VAD regulations should ensure that specific population sub-groups are neither over-represented in assisted suicide cases nor denied fair and equitable access [32]. Framing the debate across these different levels — individual, contextual, and structural — would support the implementation of appropriate safeguards and help build broader consensus.

## Declarations

**Ethics approval and consent to participate**: Not applicable. This research did not require ethical approval because it uses publicly available data.

**Consent for publication**: All authors have read and approved the final version of this manuscript and consent to its submission for publication.

**Availability of data and materials**: Hansards were download from the UK parliament Hansards website (https://hansard.parliament.uk)

**Competing interests**: NH is a member of the Federal Commission for the Control and Evaluation of Euthanasia (FCCEE). The FCCEE has no role in the design and making of the study and the intention to submit. JW, JGH and NH report no competing of interest.

**Funding**: This research did not receive funding.

**Authors’ contributions**: Conceptualisation: JW, NH; Methodology: JW, NH; Software: JW; Validation: NH, JGH; Formal analysis: JW; Investigation: JW, NH; Resources: JW, NH; Data Curation: NH; Writing - Original Draft; JW; Writing - Review & Editing: NH, JGH; Visualization: JW, JGH; Supervision: JW; Project administration: JW, NH; Funding acquisition: NA

## Data Availability

all data produced in the present study are available online and can be download from the UK parliament Hansards website (https://hansard.parliament.uk)

## Acknowledgements

Not applicable

