## Supplementary files for "Categorising Coercion in Assisted Dying. An AI-Assisted Analysis of the House of Commons Debate on the 2025 Terminally Ill Adults Bill in England and Wales"

### **Supplementary file S.1. Stages of the 2025 Terminally Ill Adults (End of Life) Bill in England and Wales, key outcomes, vote margins and number of recorder significant interventions**

**In England and Wales – the focus of this study – the Terminally Ill Adults (End of Life) Bill received its First Reading in the House of Commons on 16 October 2024. Labour MP Kim Leadbeater presented the Bill after winning the Private Member’s Bill ballot. The Bill then progressed to its Second Reading on 29 November 2024, during which its core principles were debated. Following a free vote, it passed with 330 votes in favour and 275 against. This approval committed the Bill to committee scrutiny. The Committee Stage took place over 29 sittings between 21 January and 25 March 2025. A 23-member Public Bill Committee conducted a line-by-line examination of the legislation. During this scrutiny period, over 500 amendments were proposed, of which 102 were ultimately adopted. Significant changes included replacing the original requirement for High Court approval with the newly created Assisted Dying Review Panels. These panels consisted of a senior legal figure, a psychiatrist and a social worker. Other key amendments mandated independent advocates for patients with disabilities or mental health conditions, required doctors to complete specialised training on detecting coercion, and established a four-year implementation period after Royal Assent. The Bill then entered its Report Stage on 16 May and 13 June 2025, where further amendments were debated by the full House of Commons. These included a ban on advertising assisted dying services and a requirement for the government to publish a national palliative care assessment within one year. However, proposals to exclude patients motivated by financial hardship were rejected.**

**The final Commons stage occurred with the Third Reading on 20 June 2025. After debate on the amended version, the Bill passed with 314 votes in favour to 291 against. This narrower margin compared to the Second Reading reflected adjustments made during scrutiny. The bill has been sent to the House of Lords for scrutiny in June 2025, expected to continue through late 2025 and 2026. Should the Lords propose further amendments, the Bill will return to the Commons for reconsideration. The Bill would apply only to England and Wales, with Scotland and the Isle of Man developing separate legislation.**

| **Stage** | **Date** | **Key Outcome** | **Vote Margin** | **Number of recorded significant interventions** |
| --- | --- | --- | --- | --- |
| First Reading | 16-Oct-24 | Bill presented to Commons | – | 32 |
| Second Reading | 29-Nov-24 | Approved for committee scrutiny | 330 (aye)–275 | 42 |
| Committee Stage: first sitting | 21-Jan-25 | After the bill has passed second reading, it moved on to the committee stage, where it was examined in detail. The committee comprised 23 members, balanced to reflect both the political makeup of the Commons and the range of views on this sensitive issue. The group included 14 MPs broadly supportive of the bill and 9 opposed, drawn from Labour (12 members, including the bill's sponsor Kim Leadbeater and Stephen Kinnock, the Minister for Palliative Care), the Conservatives (4 members, including opponents Danny Kruger and Rebecca Paul), the Liberal Democrats (3 members), and Plaid Cymru (1 member, Liz Saville-Roberts, who supported the legislation). The committee conducted an unusually thorough examination of this Private Member's Bill, including for the first time in such proceedings the taking of oral evidence from external specialists. Medical experts provided testimony, alongside legal. Advocacy groups like Dignity in Dying contributed perspectives, as did disability rights campaigners and medical. During detailed line-by-line scrutiny lasting several weeks, committee members proposed, debated and voted on numerous amendments informed by this expert testimony. | |  |
| Committee Stage: second sitting | 28-Jan-25 |  |  |  |
| Committee Stage: third sitting | 28-Jan-25 |  |  |  |
| Committee Stage: fourth sitting | 29-Jan-25 |  |  |  |
| Committee Stage: fifth sitting | 29-Jan-25 |  |  |  |
| Committee Stage: sixty sitting | 30-Jan-25 |  |  |  |
| Committee Stage: seventh sitting | 30-Jan-25 |  |  |  |
| Committee Stage: eighth sitting | 11-Feb-25 |  |  |  |
| Committee Stage: nineth sitting | 11-Feb-25 |  |  |  |
| Committee Stage: tenth sitting | 12-Feb-25 |  |  |  |
| Committee Stage: eleventh sitting | 12-Feb-25 |  |  |  |
| Committee Stage: twelfth sitting | 25-Feb-25 |  |  |  |
| Committee Stage: thirteenth sitting | 25-Feb-25 |  |  |  |
| Committee Stage: fourteenth sitting | 26-Feb-25 |  |  |  |
| Committee Stage: fifteenth sitting | 26-Feb-25 |  |  |  |
| Committee Stage: sixteenth sitting | 04-Mar-25 |  |  |  |
| Committee Stage: seventeenth sitting | 04-Mar-25 |  |  |  |
| Committee Stage: eighteenth sitting | 05-Mar-25 |  |  |  |
| Committee Stage: nineteenth sitting | 05-Mar-25 |  |  |  |
| Committee Stage: twentieth sitting | 11-Mar-25 |  |  |  |
| Committee Stage: twenty-fifth sitting | 11-Mar-25 |  |  |  |
| Committee Stage: twenty-second sitting | 12-Mar-25 |  |  |  |
| Committee Stage: twenty-third sitting | 12-Mar-25 |  |  |  |
| Committee Stage: twenty-fourth sitting | 18-Mar-25 |  |  |  |
| Committee Stage: twenty-fifth sitting | 18-Mar-25 |  |  |  |
| Committee Stage: twenty-sixth sitting | 19-Mar-25 |  |  |  |
| Committee Stage: twenty-seventh sitting | 19-Mar-25 |  |  |  |
| Committee Stage: twenty-eighth sitting | 25-Mar-25 |  |  |  |
| Committee Stage: twenty-nineth sitting | 25-Mar-25 |  |  |  |
| Report Stage | May–Jun 2025 | Safeguard amendments adopted | Mixed votes |  |
| Third Reading | 20-Jun-25 | Final Commons approval | 314 (aye)–291 | 44 |
| House of Lords | Late 2025–2026 | Ongoing scrutiny | Pending |  |

### **Supplementary file S.2. Reading sessions statistics**

#### Supplementary file S.2.1. Number of speeches analysed, position for or against the bill and mention of coercion

| **Stage** | **Date** | **Speeches analysed** | **Position** | | **Mention of coercion** |
| --- | --- | --- | --- | --- | --- |
|  |  |  | In favour | Against |  |
| First Reading | 16-Oct-24 | 31 | 12 | 18 | 21 |
| Second Reading | 29-Nov-24 | 42 | 21 | 20 | 14 |
| Third Reading | 20-Jun-25 | 43 | 19 | 24 | 28 |
| Total Readings |  | 116 | 52 | 62 | 63 |

#### Supplementary file S.2.2. MPs, party membership, position, mention of coercion, types of coercion (exact wording)

| **Stage** | **MP name** | **Party** | **Position** | **Mentions of coercion** | **Coercion types** |
| --- | --- | --- | --- | --- | --- |
| first | Kim Leadbeater | Labour | for | TRUE | robust protections against coercion |
| first | Kim Leadbeater | Labour | for | TRUE | controlling or coercive behaviour, economic abuse, psychological, emotional or other abuse |
| first | Rebecca Paul | Con | against | TRUE | coercion from family members or friends, societal coercion and circumstances, pressure to go down the assisted dying route, concern not to be a burden, social and welfare issues such as being homeless |
| first | Dame Meg Hillier | Lab/Co-op | against | TRUE | feel pressured to end their lives, potentially unintentionally coercing that patient, feel that they are being steered in that direction, feel pressured into ending their lives sooner than they would wish to, latched on to by someone who is coercively controlling the person |
| first | Dr Neil Shastri-Hurst | Con | against | FALSE |  |
| first | Anneliese Dodds | Lab/Co-op | for | TRUE | coercion by others, gentle advice or suggestions from authority figures, school the subject of their coercion in how to respond to questioning |
| first | Sir Edward Leigh | Con | against | TRUE | pressure on the workers in that care home, substantial moral dilemma for that person, profoundly so if they are individually opposed to Assisted Dying, the pressure on the workers in that care home, deeply traumatic experiences for loved ones |
| first | Tom Rutland | Lab | for | FALSE |  |
| first | Tom Gordon | LD | for | TRUE | cruel and unnecessary barrier, intolerable suffering, forced to consider dying earlier, endure prolonged suffering, act prematurely |
| first | Florence Eshalomi | Labour | against | TRUE | coercion of minority communities, feelings of burden or coercion |
| first | Sarah Bool | Con | for | TRUE | not being coerced |
| first | Dr Tidball | Labour | for | FALSE |  |
| first | Liz Jarvis | LD | for | TRUE | coercive, controlling and economic abuse |
| first | Rachael Maskell | Lab/Co-op | against | TRUE | intrinsic or extrinsic |
| first | Sir Jeremy Wright | Con | against | TRUE | coercion or pressure from anyone else, new partner isolated the person, evidence of coercion |
| first | Kevin Bonavia | Lab | for | FALSE |  |
| first | Warinder Juss | Lab | for | FALSE |  |
| first | Damian Hinds | Con | against | TRUE | pressure from others, pressure from oneself |
| first | Cat Eccles | Lab | against | FALSE |  |
| first | Siân Berry | Green | for | TRUE | additional intrusive steps and interrogation, extra processes, compulsory psychological investigation |
| first | Naz Shah | Labour | against | TRUE | people will fall through that loophole, they would bring themselves within the scope of this Bill as it is written, If the safeguards in the Bill fail even once, it will be a young woman like Jessica who dies |
| first | Kit Malthouse | Con | for | TRUE | coercively controlled by a partner |
| first | Lizzi Collinge | Lab | against | FALSE |  |
| first | Dr Ben Spencer | Con | against | TRUE | a true and free choice cannot be made under coercion |
| first | Daniel Francis | Labour | against | TRUE | could be a burden on their elderly parents, presumed to have capacity |
| first | Carla Lockhart | DUP | against | TRUE | coercion and undue influence, risk of coercion |
| first | Dame Meg Hillier | Labour | against | FALSE |  |
| first | Dr Opher |  | against | TRUE | make it a crime, so we could be convicted for doing so |
| first | Sarah Olney | LD | against | TRUE | putting vulnerable people at risk, doctor shopping, insufficiency of the Mental Capacity Act, severe risk that the physical impact of an eating disorder can be diagnosed as a terminal illness, mental health conditions or other factors that may lead people to seek a premature end to their life |
| first | Dr Kieran Mullan | Con | against | TRUE | question the integrity, sincerity or understanding of those MPs seeking a different outcome to them, clumsy criticism of those whose objections to the Bill are thought to be centred in their religious beliefs |
| first | Stephen Kinnock | Labour | for | TRUE | no person is under any duty to participate,employees may end up with conflicting obligations,protection for employees from detriment and unfair dismissal by their employers should they choose to participate in the provision of assisted dying,If employers can prevent their entire workforce from participating in the provision of assisted dying, the service might not be available or could be much more difficult to access,require a person to leave the care home in which they reside to access assistance under the Bill,a person who is terminally ill could be asked or required to leave that care home or hospice to receive assistance under the Bill |
| second | Kim Leadbeater | Lab | for | FALSE |  |
| second | Sarah Olney | LD | against | TRUE | if they suffer from a mental illness or eating disorder, if they are from a low-income or ethnic minority background, if they suffer from a disability or if they are in an abusive relationship |
| second | Dr Jeevun Sandher | Lab | against | FALSE |  |
| second | Rebecca Smith | Con | against | TRUE | coercion or abuse, particularly where family members have conflicting financial, social or emotional burdens |
| second | Paul Waugh | Lab/Co-op | for | TRUE | self-coercion, coercive control |
| second | Dame Caroline Dinenage | Con | for | FALSE |  |
| second | Sadik Al-Hassan | Lab | for | FALSE |  |
| second | Dame Harriett Baldwin | Con | against | FALSE |  |
| second | Alison Hume | Lab | for | FALSE |  |
| second | Gregory Stafford | Conservative | against | FALSE |  |
| second | Adam Jogee | Lab | against | FALSE |  |
| second | Tom Gordon | LD | against | TRUE | pushed to taking decisions that they should not be |
| second | Tony Vaughan | Lab | for | TRUE | coercion or fraudulent practice |
| second | John Glen | Con | for | FALSE |  |
| second | Dr Allison Gardner | Lab | for | TRUE | gaming of the system, doctor shopping |
| second | Mr Paul Kohler | LD | against | TRUE | safeguards against coercion in all its forms, assessing coercive control |
| second | Rachel Taylor | Labour | for | FALSE |  |
| second | Dr Ben Spencer | Conservative | against | TRUE | doctors’ duties under article 2 of the European convention on human rights, the NHS’s clinical duties around suicide prevention, duties relating to the Mental Health Act 1983 |
| second | Marsha De Cordova | Lab | against | TRUE | feel subtly pressured to end their lives due to social attitudes and lack of appropriate services and support |
| second | Ellie Chowns | Green | for | FALSE |  |
| second | Patricia Ferguson | Labour | for | FALSE |  |
| second | Dr Caroline Johnson | Con | against | FALSE |  |
| second | Alex Barros-Curtis | Lab | for | FALSE |  |
| second | Robin Swann | UUP | against | TRUE | ministerial diktat, Henry VIII regulations |
| second | Blair McDougall | Lab | for | TRUE | economic coercion, social and cultural context, economic disadvantage, financial motivations, societal and cultural pressure |
| second | Simon Hoare | Conservative | for | FALSE |  |
| second | Catherine Fookes | Lab | for | FALSE |  |
| second | Liz Saville Roberts | PC | for | TRUE | questions of coercion and capacity, potential questions of coercion and capacity |
| second | Dame Siobhain McDonagh | Lab | against | FALSE |  |
| second | Calum Miller | LD | for | FALSE |  |
| second | Dr Rupa Huq | Lab | against | TRUE | coercion, duress, the billionaire price of London property, elder abuse, relatives wanting to speed up granny or grandad’s probate, people could convince themselves that elderly relatives would be better off out of the way |
| second | Richard Tice | Reform | for | FALSE |  |
| second | Melanie Ward | Lab | for | TRUE | fear of being a burden on their family, friends or caregivers, pressure from others who do not have their best interests at heart |
| second | Munira Wilson | LD | against | TRUE | less likely to access the healthcare services they deserve, less likely to access assisted dying, do not have the loudest voices or well-funded campaigns to support them, people will not have a genuine choice at the end of their life |
| second | Lloyd Hatton | Lab | for | FALSE |  |
| second | Carla Lockhart | DUP | against | FALSE |  |
| second | John Grady | Lab | against | TRUE | coercive control, domestic abuse, elder abuse, societal pressures |
| second | Sir John Hayes | Conservative | against | TRUE | encouraged, perhaps even forced, to take a decision, felt they were a burden to their family |
| second | Rachael Maskell | Labour | against | FALSE |  |
| second | Dr Kieran Mullan | Con |  | FALSE |  |
| second | Stephen Kinnock | Labour |  | FALSE |  |
| third | Kim Leadbeater | Lab | for | TRUE | coercive control, coercion, dishonesty or pressure |
| third | Sir James Cleverly | Con | against | TRUE | the pressure that individuals put on themselves, coercion, on the pressure that individuals put on themselves and on medical professionals raising the issue |
| third | Diane Abbott | Lab | against | TRUE | silence: it is the failure to answer when a question is put, coercion in assisted dying, foul play, lack of agency in a family context, lack of access to good palliative care |
| third | Mark Garnier | Con | for | FALSE |  |
| third | Naz Shah | Lab | against | TRUE | people with anorexia, those with mental health conditions, troubled people—those who would be vulnerable, the most vulnerable, those who have faced the greatest hardships in life and feel like a burden, those who feel like giving up |
| third | Josh Babarinde | LD | for | TRUE | coerce a relative in such a way, Self-coercion |
| third | Peter Prinsley | Lab | for | FALSE |  |
| third | Sarah Olney | LD | against | FALSE |  |
| third | Vicky Foxcroft | Lab | against | TRUE | do not resuscitate notices unilaterally pasted on their medical records, without them being informed, susceptible to coercion |
| third | Sir Iain Duncan Smith | Con | against | FALSE |  |
| third | Rachel Hopkins | Lab | for | TRUE | coercion and capacity |
| third | Sir Edward Leigh | Con | against | FALSE |  |
| third | Andy Slaughter | Lab | for | TRUE | the risk of coercion, putting a caring loved one at risk of prosecution |
| third | Mike Wood | Con | against | TRUE | wish to avoid being a burden, reading too much into the doctor’s suggestion when they raised assisted death as something to consider |
| third | David Smith | Lab | against | TRUE | coercion, pressure, subtly pushing the option of assisted death |
| third | Kit Malthouse | Con | for | TRUE | forced to travel abroad to die in lonely circumstances, prosecuted for holding the hand of someone they have loved for 50 years, lonely suicides in quiet suburban bedrooms |
| third | Kevin McKenna | Lab | for | TRUE | well-meaning families and clinicians provide a degree of coercion |
| third | Gavin Robinson | DUP | against | TRUE | their trade unions and their governing bodies do not support them or insure them in that endeavour |
| third | Dr Beccy Cooper | Lab | for | TRUE | coerced into taking a decision to end their life, coercion in this space, coerced into doing so by a person or persons with malign intent |
| third | Tom Tugendhat | Con | against | TRUE | intimidated, fear, neglect, verbal abuse, denial of essential care, devalued, suggested to end life, pressure |
| third | Jen Craft | Lab | against | TRUE | coerced or given bad advice |
| third | Luke Taylor | LD | for | FALSE |  |
| third | Maureen Burke | Lab | for | FALSE |  |
| third | Sir Jeremy Wright | Con | against | TRUE | the fear of prosecution for acts of love and mercy, is encouraged in the belief that their life is not valuable and valued to their very last moment |
| third | Preet Kaur Gill | Lab/Co-op | against | TRUE | could not compel them to attend, would not be required to question witnesses, would not give evidence under oath, nobody would cross-examine them, palliative care provision is woefully inadequate, the worst served are also the most disadvantaged, the provision of palliative care is likely to be compromised |
| third | Mr Peter Bedford | Con | for | FALSE |  |
| third | Dame Chi Onwurah | Lab | against | TRUE | the state to take the life of a citizen, private companies to kill private citizens, powerful economic and personal incentives for both the state and family members to encourage the vulnerable into taking their own lives |
| third | Nigel Huddleston | Con | against | TRUE | the vulnerable, who are exposed to the risk of coercion |
| third | David Burton-Sampson | Lab | for | TRUE | safeguards in place to ensure that an individual is not coerced |
| third | Wendy Morton | Con | against | FALSE |  |
| third | John McDonnell | Ind | for | TRUE | Hide away the drugs over a period of time, send their families away, and then take the drugs and die a lonely death, starved themselves to death because there was no other option |
| third | Munira Wilson | LD | against | TRUE | people choosing to end their life before they want to, choosing not to access the care that they need |
| third | Jess Asato | Lab | against | TRUE | coercion and abuse, poverty, the patriarchy, racism, trauma, ill health, state and societal failure, financial abuse, cuckooing, feeling like a burden |
| third | Sarah Green | LD | for | TRUE | interviewed by the police for supporting her husband in going to Dignitas, faced the uncertainty of a police investigation |
| third | Paula Barker | Lab | for | TRUE | guard against coercion,detecting coercion |
| third | Dr Ben Spencer | Con | against | TRUE | those who feel a burden, those who are made to feel a burden, those who are abused |
| third | Matthew Patrick | Lab | against | TRUE | quiet, imperceptible and unspoken coercion, vulnerable people who would feel a duty to die |
| third | Christine Jardine | LD | for | FALSE |  |
| third | Lewis Atkinson | Lab | for | TRUE | no checks on capacity or coercion |
| third | Dr Neil Hudson | Con | against | TRUE | pressure to proceed down this path, possible pressure that the Bill will place on medical practitioners |
| third | Lola McEvoy | Lab | against | FALSE |  |
| third | Dr Kieran Mullan | Con | against | FALSE |  |
| third | Stephen Kinnock | Labour | for | FALSE |  |

#### Supplementary file S.2.3. Coercion types by speaker

| **Speaker name** | **Party** | **Position** | **Coercion types** | | | | | | | **HC** |
| --- | --- | --- | --- | --- | --- | --- | --- | --- | --- | --- |
|  |  |  | ***family*** | ***medical*** | ***poor care*** | ***economic*** | ***ethnic*** | ***self*** | ***mental*** |  |
| Kim Leadbeater | Labour | for | 0 | 0 | 0 | 0 | 0 | 0 | 0 | first |
| Kim Leadbeater | Labour | for | 0 | 0 | 0 | 0 | 0 | 0 | 0 | first |
| Rebecca Paul | Con | against | 1 | 1 | 1 | 1 | 0 | 1 | 1 | first |
| Dame Meg Hillier | Lab/Co-op | against | 1 | 1 | 1 | 0 | 0 | 1 | 1 | first |
| Dr Neil Shastri-Hurst | Con | against | 0 | 0 | 0 | 0 | 0 | 0 | 0 | first |
| Anneliese Dodds | Lab/Co-op | for | 1 | 1 | 1 | 1 | 1 | 1 | 1 | first |
| Sir Edward Leigh | Con | against | 0 | 0 | 1 | 1 | 0 | 0 | 0 | first |
| Tom Rutland | Lab | for | 0 | 0 | 0 | 0 | 0 | 0 | 0 | first |
| Tom Gordon | LD | for | 0 | 0 | 0 | 0 | 0 | 0 | 0 | first |
| Florence Eshalomi | Labour | against | 0 | 0 | 0 | 0 | 1 | 0 | 0 | first |
| Sarah Bool | Con | against | 0 | 0 | 0 | 0 | 0 | 0 | 0 | first |
| Dr Tidball | Labour | for | 0 | 0 | 0 | 0 | 0 | 0 | 0 | first |
| Liz Jarvis | LD | for | 0 | 0 | 0 | 0 | 0 | 0 | 0 | first |
| Rachael Maskell | Lab/Co-op | against | 0 | 0 | 1 | 0 | 0 | 1 | 1 | first |
| Sir Jeremy Wright | Con | against | 1 | 0 | 0 | 0 | 0 | 0 | 0 | first |
| Kevin Bonavia | Lab | for | 0 | 0 | 0 | 0 | 0 | 0 | 0 | first |
| Warinder Juss | Lab | for | 0 | 0 | 0 | 0 | 0 | 0 | 0 | first |
| Damian Hinds | Con | against | 1 | 1 | 1 | 1 | 0 | 1 | 1 | first |
| Cat Eccles | Lab | against | 0 | 0 | 1 | 0 | 0 | 0 | 0 | first |
| Siân Berry | Green | for | 0 | 0 | 0 | 0 | 0 | 0 | 0 | first |
| Naz Shah | Labour | against | 0 | 0 | 0 | 0 | 0 | 0 | 0 | first |
| Kit Malthouse | Con | for | 0 | 0 | 0 | 0 | 0 | 0 | 0 | first |
| Lizzi Collinge | Lab | against | 0 | 0 | 0 | 0 | 0 | 0 | 0 | first |
| Dr Ben Spencer | Con | against | 0 | 0 | 1 | 0 | 0 | 0 | 1 | first |
| Daniel Francis | Labour | against | 1 | 0 | 0 | 0 | 0 | 0 | 1 | first |
| Carla Lockhart | DUP | against | 0 | 1 | 1 | 0 | 0 | 0 | 1 | first |
| Dame Meg Hillier | Labour | against | 0 | 0 | 0 | 0 | 0 | 0 | 0 | first |
| Dr Opher |  | against | 0 | 0 | 0 | 0 | 0 | 0 | 0 | first |
| Sarah Olney | LD | against | 0 | 0 | 0 | 0 | 0 | 0 | 1 | first |
| Dr Kieran Mullan | Con |  | 0 | 0 | 0 | 0 | 0 | 0 | 0 | first |
| Stephen Kinnock | Labour | for | 0 | 0 | 0 | 0 | 0 | 0 | 0 | first |
| Kim Leadbeater | Lab | for | 0 | 0 | 0 | 0 | 0 | 0 | 0 | second |
| Sarah Olney | LD | against | 0 | 0 | 0 | 1 | 1 | 0 | 1 | second |
| Dr Jeevun Sandher | Lab | against | 0 | 0 | 0 | 0 | 0 | 0 | 0 | second |
| Rebecca Smith | Con | against | 1 | 1 | 1 | 1 | 1 | 1 | 1 | second |
| Paul Waugh | Lab/Co-op | for | 0 | 0 | 0 | 0 | 0 | 1 | 0 | second |
| Dame Caroline Dinenage | Con | for | 0 | 0 | 0 | 0 | 0 | 0 | 0 | second |
| Sadik Al-Hassan | Lab | for | 0 | 0 | 0 | 0 | 0 | 0 | 0 | second |
| Dame Harriett Baldwin | Con | against | 0 | 0 | 0 | 0 | 0 | 0 | 0 | second |
| Alison Hume | Lab | for | 0 | 0 | 0 | 0 | 0 | 0 | 0 | second |
| Gregory Stafford | Conservative | against | 0 | 0 | 0 | 0 | 0 | 0 | 0 | second |
| Adam Jogee | Lab | against | 0 | 0 | 0 | 0 | 0 | 0 | 0 | second |
| Tom Gordon | LD | for | 0 | 0 | 0 | 0 | 0 | 0 | 0 | second |
| Tony Vaughan | Lab | for | 0 | 0 | 0 | 0 | 0 | 0 | 0 | second |
| John Glen | Con | against | 0 | 0 | 0 | 0 | 0 | 0 | 0 | second |
| Dr Allison Gardner | Lab | against | 0 | 0 | 0 | 0 | 0 | 0 | 0 | second |
| Paul Kohler | LD | against | 0 | 0 | 1 | 0 | 0 | 0 | 0 | second |
| Rachel Taylor | Labour | for | 0 | 0 | 0 | 0 | 0 | 0 | 0 | second |
| Dr Ben Spencer | Conservative | against | 0 | 0 | 0 | 0 | 0 | 0 | 1 | second |
| Marsha De Cordova | Lab | against | 1 | 1 | 1 | 1 | 0 | 0 | 1 | second |
| Ellie Chowns | Green | for | 0 | 0 | 0 | 0 | 0 | 0 | 0 | second |
| Patricia Ferguson | Labour | for | 0 | 0 | 0 | 0 | 0 | 0 | 0 | second |
| Dr Caroline Johnson | Con | against | 0 | 0 | 0 | 0 | 0 | 0 | 0 | second |
| Alex Barros-Curtis | Lab | for | 0 | 0 | 0 | 0 | 0 | 0 | 0 | second |
| Robin Swann | UUP | against | 0 | 0 | 0 | 0 | 0 | 0 | 0 | second |
| Blair McDougall | Lab | against | 0 | 0 | 1 | 1 | 0 | 0 | 0 | second |
| Simon Hoare | Conservative | for | 0 | 0 | 0 | 0 | 0 | 0 | 0 | second |
| Catherine Fookes | Lab | for | 0 | 0 | 0 | 0 | 0 | 0 | 0 | second |
| Liz Saville Roberts | PC | for | 0 | 0 | 0 | 0 | 0 | 0 | 0 | second |
| Dame Siobhain McDonagh | Lab | against | 0 | 0 | 0 | 0 | 0 | 0 | 0 | second |
| Calum Miller | LD | for | 0 | 0 | 0 | 0 | 0 | 0 | 0 | second |
| Dr Rupa Huq | Lab | against | 1 | 1 | 1 | 1 | 1 | 0 | 1 | second |
| Richard Tice | Reform | for | 0 | 0 | 0 | 0 | 0 | 0 | 0 | second |
| Melanie Ward | Lab | against | 1 | 1 | 1 | 1 | 0 | 1 | 1 | second |
| Munira Wilson | LD | against | 0 | 0 | 1 | 1 | 1 | 0 | 0 | second |
| Lloyd Hatton | Lab | for | 0 | 0 | 0 | 0 | 0 | 0 | 0 | second |
| Carla Lockhart | DUP | against | 0 | 0 | 0 | 0 | 0 | 0 | 0 | second |
| John Grady | Lab | against | 0 | 0 | 1 | 1 | 0 | 1 | 1 | second |
| Sir John Hayes | Conservative | against | 1 | 1 | 0 | 1 | 0 | 1 | 1 | second |
| Rachael Maskell | Labour | against | 0 | 0 | 0 | 0 | 0 | 0 | 0 | second |
| Dr Kieran Mullan | Con | against | 0 | 0 | 0 | 0 | 0 | 0 | 0 | second |
| Stephen Kinnock | Labour | for | 0 | 0 | 0 | 0 | 0 | 0 | 0 | second |
| Kim Leadbeater | Lab | for | 0 | 0 | 0 | 0 | 0 | 0 | 0 | third |
| Sir James Cleverly | Con | against | 0 | 0 | 0 | 0 | 0 | 0 | 0 | third |
| Diane Abbott | Lab | against | 1 | 0 | 1 | 0 | 1 | 0 | 1 | third |
| Mark Garnier | Con | for | 0 | 0 | 0 | 0 | 0 | 0 | 0 | third |
| Naz Shah | Lab | against | 0 | 0 | 0 | 0 | 0 | 0 | 1 | third |
| Josh Babarinde | LD | for | 0 | 0 | 0 | 0 | 0 | 1 | 0 | third |
| Peter Prinsley | Lab | for | 0 | 0 | 0 | 0 | 0 | 0 | 0 | third |
| Sarah Olney | LD | against | 0 | 0 | 0 | 0 | 0 | 0 | 0 | third |
| Vicky Foxcroft | Lab | against | 1 | 1 | 1 | 1 | 0 | 1 | 1 | third |
| Sir Iain Duncan Smith | Con | against | 0 | 0 | 1 | 0 | 0 | 0 | 0 | third |
| Rachel Hopkins | Lab | for | 0 | 0 | 0 | 0 | 0 | 0 | 0 | third |
| Sir Edward Leigh | Con | against | 0 | 0 | 0 | 0 | 0 | 0 | 0 | third |
| Andy Slaughter | Lab | for | 0 | 0 | 0 | 0 | 0 | 0 | 0 | third |
| Mike Wood | Con | against | 1 | 1 | 0 | 0 | 0 | 1 | 0 | third |
| David Smith | Lab | against | 1 | 0 | 0 | 1 | 0 | 1 | 0 | third |
| Kit Malthouse | Con | for | 0 | 0 | 0 | 0 | 0 | 0 | 0 | third |
| Kevin McKenna | Lab | for | 1 | 1 | 0 | 0 | 0 | 0 | 0 | third |
| Gavin Robinson | DUP | against | 0 | 0 | 0 | 0 | 0 | 0 | 0 | third |
| Dr Beccy Cooper | Lab | for | 0 | 0 | 0 | 0 | 0 | 0 | 0 | third |
| Tom Tugendhat | Con | against | 0 | 1 | 1 | 0 | 0 | 0 | 1 | third |
| Jen Craft | Lab | against | 1 | 1 | 1 | 0 | 0 | 0 | 1 | third |
| Luke Taylor | LD | for | 0 | 0 | 0 | 0 | 0 | 0 | 0 | third |
| Maureen Burke | Lab | for | 0 | 0 | 0 | 0 | 0 | 0 | 0 | third |
| Sir Jeremy Wright | Con | against | 0 | 0 | 0 | 0 | 0 | 0 | 0 | third |
| Preet Kaur Gill | Lab/Co-op | against | 1 | 1 | 1 | 1 | 1 | 0 | 1 | third |
| Peter Bedford | Con | for | 0 | 0 | 0 | 0 | 0 | 0 | 0 | third |
| Dame Chi Onwurah | Lab | against | 1 | 0 | 0 | 1 | 1 | 0 | 1 | third |
| Nigel Huddleston | Con | against | 0 | 0 | 0 | 0 | 0 | 0 | 0 | third |
| David Burton-Sampson | Lab | for | 0 | 0 | 1 | 0 | 0 | 0 | 0 | third |
| Wendy Morton | Con | against | 0 | 0 | 0 | 0 | 0 | 0 | 0 | third |
| John McDonnell | Ind | for | 0 | 0 | 0 | 0 | 0 | 0 | 0 | third |
| Munira Wilson | LD | against | 0 | 0 | 1 | 0 | 1 | 0 | 0 | third |
| Jess Asato | Lab | against | 1 | 1 | 1 | 1 | 0 | 1 | 1 | third |
| Sarah Green | LD | for | 0 | 0 | 0 | 0 | 0 | 0 | 0 | third |
| Paula Barker | Lab | for | 0 | 0 | 0 | 0 | 0 | 0 | 0 | third |
| Dr Ben Spencer | Con | against | 0 | 0 | 1 | 0 | 0 | 1 | 1 | third |
| Matthew Patrick | Lab | against | 0 | 0 | 1 | 0 | 1 | 1 | 0 | third |
| Christine Jardine | LD | for | 0 | 0 | 0 | 0 | 0 | 0 | 0 | third |
| Lewis Atkinson | Lab | for | 0 | 0 | 0 | 0 | 0 | 0 | 0 | third |
| Dr Neil Hudson | Con | against | 1 | 1 | 1 | 0 | 0 | 0 | 0 | third |
| Lola McEvoy | Lab | against | 0 | 0 | 1 | 0 | 0 | 0 | 0 | third |
| Dr Kieran Mullan | Con | for | 0 | 0 | 0 | 0 | 0 | 0 | 0 | third |
| Stephen Kinnock | Labour | for | 0 | 0 | 0 | 0 | 0 | 0 | 0 | third |
| Total |  |  | 21 | 18 | 30 | 18 | 11 | 17 | 26 |  |

### **Supplementary file S.3. Sitting sessions statistics**

#### Supplementary file S.3.1. Number of speeches analysed per sitting and mention of coercion types

| **Sitting** | **speeches** | **speakers** | **Coercion types** | | | | | | |
| --- | --- | --- | --- | --- | --- | --- | --- | --- | --- |
|  |  |  | ***family*** | ***medical*** | ***poor care*** | ***economic*** | ***ethnic*** | ***self*** | ***mental*** |
| 0 | 200 | 104 | 19 | 4 | 14 | 13 | 2 | 8 | 12 |
| 1 | 76 | 22 | 0 | 0 | 0 | 0 | 0 | 0 | 2 |
| 2 | 192 | 35 | 3 | 1 | 3 | 2 | 1 | 0 | 4 |
| 3 | 259 | 48 | 7 | 1 | 5 | 2 | 2 | 4 | 4 |
| 4 | 156 | 40 | 1 | 2 | 2 | 1 | 1 | 1 | 4 |
| 5 | 216 | 41 | 4 | 5 | 8 | 8 | 7 | 4 | 7 |
| 6 | 66 | 23 | 1 | 1 | 1 | 0 | 0 | 1 | 2 |
| 7 | 252 | 52 | 5 | 4 | 3 | 1 | 2 | 2 | 10 |
| 8 | 140 | 28 | 1 | 1 | 0 | 0 | 0 | 0 | 10 |
| 9 | 175 | 30 | 2 | 4 | 4 | 1 | 0 | 4 | 12 |
| 10 | 129 | 27 | 8 | 2 | 1 | 0 | 0 | 1 | 2 |
| 11 | 193 | 35 | 6 | 2 | 2 | 7 | 1 | 5 | 3 |
| 12 | 112 | 23 | 0 | 1 | 4 | 2 | 2 | 1 | 5 |
| 13 | 140 | 28 | 0 | 4 | 1 | 1 | 0 | 0 | 8 |
| 14 | 86 | 19 | 1 | 3 | 1 | 1 | 0 | 1 | 8 |
| 15 | 142 | 28 | 3 | 5 | 2 | 1 | 2 | 3 | 8 |
| 16 | 90 | 22 | 0 | 2 | 0 | 0 | 0 | 0 | 1 |
| 17 | 148 | 21 | 1 | 2 | 0 | 1 | 3 | 1 | 5 |
| 18 | 93 | 19 | 1 | 1 | 0 | 2 | 1 | 1 | 4 |
| 19 | 213 | 23 | 0 | 3 | 0 | 0 | 1 | 0 | 11 |
| 20 | 87 | 20 | 1 | 0 | 0 | 2 | 0 | 1 | 3 |
| 21 | 216 | 31 | 6 | 1 | 1 | 1 | 0 | 2 | 3 |
| 22 | 75 | 16 | 8 | 1 | 2 | 2 | 0 | 3 | 4 |
| 23 | 189 | 26 | 2 | 1 | 1 | 3 | 1 | 3 | 4 |
| 24 | 95 | 16 | 3 | 1 | 1 | 0 | 0 | 3 | 3 |
| 25 | 303 | 27 | 1 | 1 | 1 | 1 | 1 | 1 | 0 |
| 26 | 65 | 18 | 0 | 0 | 3 | 0 | 0 | 0 | 0 |
| 27 | 212 | 25 | 5 | 0 | 0 | 1 | 3 | 1 | 1 |
| 28 | 76 | 19 | 1 | 2 | 2 | 4 | 1 | 1 | 1 |
| 29 | 261 | 25 | 7 | 1 | 1 | 5 | 2 | 1 | 3 |
| Sum | 4,657 |  | 97 | 56 | 63 | 62 | 33 | 53 | 144 |

#### Supplementary file S.3.2. Coercion types by speaker

| **Speaker** | **Party** | **speeches** | **Coercion type** | | | | | | | **Sitting** |
| --- | --- | --- | --- | --- | --- | --- | --- | --- | --- | --- |
|  |  |  | ***family*** | ***medical*** | ***poor care*** | ***economic*** | ***ethnic*** | ***self*** | ***mental*** |  |
| Alicia Kearns | Con | 2 | 1 | 0 | 0 | 0 | 0 | 0 | 0 | 0 |
| Alistair Strathern | Lab | 1 | 0 | 0 | 0 | 0 | 0 | 0 | 0 | 0 |
| Andrew George | LD | 1 | 0 | 0 | 0 | 0 | 0 | 0 | 0 | 0 |
| Andy Slaughter | Lab | 3 | 0 | 0 | 0 | 0 | 0 | 0 | 0 | 0 |
| Anna Dixon | Lab | 1 | 1 | 0 | 1 | 1 | 0 | 0 | 0 | 0 |
| Barry Gardiner | Lab | 2 | 1 | 0 | 0 | 1 | 0 | 0 | 0 | 0 |
| Blair McDougall | | 1 | 0 | 1 | 0 | 0 | 0 | 0 | 1 | 0 |
| Carla Lockhart | DUP | 1 | 0 | 0 | 1 | 0 | 0 | 0 | 0 | 0 |
| Caroline Nokes | Con | 3 | 0 | 0 | 0 | 0 | 0 | 0 | 0 | 0 |
| Cat Eccles | Lab | 1 | 0 | 0 | 0 | 0 | 0 | 0 | 0 | 0 |
| Catherine Atkinson | | 1 | 0 | 0 | 0 | 0 | 0 | 0 | 0 | 0 |
| Catherine Fookes | | 1 | 0 | 0 | 0 | 0 | 0 | 0 | 0 | 0 |
| Christine Jardine | LD | 1 | 0 | 0 | 0 | 0 | 0 | 0 | 0 | 0 |
| Daisy Cooper | LD | 1 | 0 | 0 | 0 | 0 | 0 | 0 | 0 | 0 |
| Meg Hillier | Lab | 3 | 0 | 0 | 0 | 0 | 0 | 0 | 0 | 0 |
| Danny Kruger | Con | 20 | 1 | 1 | 1 | 0 | 0 | 1 | 1 | 0 |
| Danny Kruger | Con | 1 | 0 | 0 | 0 | 0 | 0 | 0 | 0 | 0 |
| David Davis | Con | 1 | 0 | 0 | 0 | 0 | 0 | 0 | 0 | 0 |
| David Davis | Con | 1 | 0 | 0 | 0 | 0 | 0 | 0 | 0 | 0 |
| Dawn Butler | Lab | 1 | 0 | 0 | 0 | 0 | 0 | 0 | 0 | 0 |
| Arthur |  | 2 | 0 | 0 | 0 | 0 | 0 | 0 | 0 | 0 |
| Ben Spencer | Con | 1 | 0 | 0 | 1 | 0 | 0 | 0 | 0 | 0 |
| Kieran Mullan | Con | 1 | 0 | 0 | 0 | 0 | 0 | 0 | 0 | 0 |
| Luke Evans | Con | 1 | 0 | 0 | 0 | 0 | 0 | 0 | 0 | 0 |
| Marie Tidball | Lab | 1 | 0 | 0 | 0 | 0 | 0 | 0 | 0 | 0 |
| Neil Shastri-Hurst | Con | 1 | 0 | 0 | 0 | 0 | 0 | 0 | 0 | 0 |
| Opher |  | 1 | 0 | 0 | 0 | 0 | 0 | 0 | 0 | 0 |
| Scott Arthur | Lab | 1 | 0 | 0 | 0 | 0 | 0 | 0 | 0 | 0 |
| Simon Opher |  | 1 | 0 | 0 | 0 | 0 | 0 | 1 | 0 | 0 |
| Spencer |  | 1 | 1 | 0 | 0 | 1 | 0 | 0 | 1 | 0 |
| Florence Eshalomi | Lab | 1 | 1 | 0 | 1 | 0 | 1 | 0 | 0 | 0 |
| Gavin Robinson | DUP | 1 | 0 | 0 | 0 | 0 | 0 | 0 | 0 | 0 |
| Gideon Amos | LD | 1 | 0 | 0 | 0 | 0 | 0 | 0 | 0 | 0 |
| Gideon Amos | LD | 1 | 0 | 0 | 0 | 0 | 0 | 0 | 0 | 0 |
| Graham Stuart | Con | 1 | 0 | 0 | 0 | 0 | 0 | 0 | 0 | 0 |
| Imran Hussain | Lab | 1 | 0 | 0 | 0 | 0 | 0 | 0 | 0 | 0 |
| Jake Richards |  | 2 | 0 | 0 | 0 | 0 | 0 | 0 | 0 | 0 |
| James Frith |  | 1 | 0 | 0 | 0 | 0 | 0 | 0 | 0 | 0 |
| Jess Asato | Lab | 1 | 1 | 0 | 0 | 0 | 0 | 0 | 1 | 0 |
| Jim Allister | TUV | 2 | 1 | 0 | 0 | 0 | 0 | 1 | 1 | 0 |
| Jim Shannon | DUP | 1 | 0 | 0 | 0 | 0 | 0 | 0 | 0 | 0 |
| Joe Robertson | Con | 1 | 0 | 0 | 0 | 0 | 0 | 0 | 0 | 0 |
| Jonathan Davies | | 1 | 0 | 0 | 0 | 0 | 0 | 0 | 0 | 0 |
| Jonathan Davies | Lab | 1 | 0 | 0 | 0 | 0 | 0 | 0 | 0 | 0 |
| Kevin McKenna | | 3 | 0 | 0 | 0 | 0 | 0 | 0 | 0 | 0 |
| Kim Leadbeater | Lab | 22 | 1 | 0 | 0 | 0 | 0 | 1 | 0 | 0 |
| Kit Malthouse | Con | 4 | 0 | 0 | 0 | 0 | 0 | 0 | 0 | 0 |
| Layla Moran | LD | 4 | 0 | 0 | 0 | 0 | 0 | 0 | 0 | 0 |
| Lewis Atkinson | Lab | 3 | 0 | 0 | 0 | 0 | 0 | 0 | 0 | 0 |
| Liz Saville Roberts | PC | 1 | 1 | 1 | 1 | 0 | 0 | 0 | 0 | 0 |
| Lizzi Collinge |  | 1 | 0 | 0 | 0 | 0 | 0 | 0 | 0 | 0 |
| Lloyd Hatton |  | 1 | 0 | 0 | 0 | 0 | 0 | 0 | 0 | 0 |
| Lola McEvoy |  | 2 | 0 | 0 | 0 | 0 | 0 | 0 | 0 | 0 |
| Madam Deputy Speaker | | 2 | 0 | 0 | 0 | 0 | 0 | 0 | 0 | 0 |
| Madam Deputy Speaker | | 3 | 0 | 0 | 0 | 0 | 0 | 0 | 0 | 0 |
| Mark Pritchard | Con | 1 | 0 | 0 | 0 | 0 | 0 | 0 | 0 | 0 |
| Mary Kelly Foy | Lab | 2 | 0 | 0 | 0 | 0 | 0 | 0 | 0 | 0 |
| Melanie Ward |  | 3 | 1 | 0 | 0 | 0 | 0 | 0 | 0 | 0 |
| Adnan Hussain | | 1 | 0 | 0 | 0 | 0 | 0 | 0 | 0 | 0 |
| Andrew Mitchell | Con | 1 | 0 | 0 | 0 | 0 | 0 | 0 | 0 | 0 |
| Bedford |  | 1 | 0 | 0 | 0 | 0 | 0 | 0 | 0 | 0 |
| Cleverly |  | 1 | 0 | 0 | 0 | 0 | 0 | 0 | 0 | 0 |
| Frith |  | 1 | 0 | 0 | 1 | 0 | 0 | 0 | 0 | 0 |
| James Cleverly | Con | 1 | 0 | 0 | 0 | 0 | 0 | 0 | 0 | 0 |
| Lee Dillon |  | 1 | 0 | 0 | 0 | 0 | 0 | 0 | 0 | 0 |
| Mitchell | Con | 1 | 0 | 0 | 0 | 0 | 0 | 0 | 0 | 0 |
| Perkins |  | 1 | 0 | 0 | 0 | 0 | 0 | 0 | 0 | 0 |
| Peter Bedford |  | 1 | 0 | 0 | 0 | 0 | 0 | 0 | 0 | 0 |
| Speaker |  | 7 | 0 | 0 | 0 | 0 | 0 | 0 | 0 | 0 |
| Toby Perkins | Lab | 1 | 0 | 0 | 0 | 0 | 0 | 0 | 0 | 0 |
| Abbott | Lab | 5 | 1 | 0 | 0 | 1 | 0 | 1 | 1 | 0 |
| Diane Abbott | Lab | 1 | 0 | 0 | 0 | 0 | 0 | 0 | 0 | 0 |
| Naz Shah | Lab | 1 | 0 | 0 | 0 | 0 | 0 | 0 | 0 | 0 |
| Neil O'Brien | Con | 1 | 0 | 0 | 0 | 0 | 0 | 0 | 0 | 0 |
| Neil O’Brien | Con | 2 | 0 | 0 | 0 | 0 | 0 | 0 | 0 | 0 |
| Paula Barker | Lab | 3 | 0 | 0 | 0 | 1 | 0 | 0 | 0 | 0 |
| Paulette Hamilton | Lab | 1 | 1 | 1 | 1 | 1 | 0 | 0 | 1 | 0 |
| Peter Prinsley |  | 3 | 0 | 0 | 0 | 0 | 0 | 0 | 0 | 0 |
| Rachael Maskell | Lab | 3 | 1 | 0 | 1 | 1 | 1 | 1 | 1 | 0 |
| Rachel Hopkins | Lab | 2 | 0 | 0 | 0 | 1 | 0 | 0 | 0 | 0 |
| Rachel Taylor |  | 1 | 0 | 0 | 0 | 0 | 0 | 0 | 0 | 0 |
| Richard Burgon | Lab | 1 | 1 | 0 | 1 | 1 | 0 | 0 | 0 | 0 |
| Richard Tice | Reform | 1 | 0 | 0 | 0 | 0 | 0 | 0 | 0 | 0 |
| Robert Jenrick | Con | 1 | 1 | 0 | 0 | 1 | 0 | 0 | 1 | 0 |
| Rosie Wrighting | | 1 | 0 | 0 | 0 | 1 | 0 | 0 | 0 | 0 |
| Ruth Jones | Lab | 1 | 0 | 0 | 0 | 0 | 0 | 0 | 0 | 0 |
| Sam Rushworth | Lab | 1 | 0 | 0 | 0 | 0 | 0 | 0 | 0 | 0 |
| Saqib Bhatti | Con | 2 | 0 | 0 | 0 | 0 | 0 | 0 | 1 | 0 |
| Shockat Adam |  | 1 | 0 | 0 | 0 | 0 | 0 | 0 | 0 | 0 |
| Simon Hoare | Con | 2 | 1 | 0 | 0 | 0 | 0 | 0 | 0 | 0 |
| Edward Leigh | Con | 2 | 0 | 0 | 1 | 0 | 0 | 0 | 0 | 0 |
| John Hayes | Con | 2 | 0 | 0 | 0 | 0 | 0 | 0 | 0 | 0 |
| Julian Lewis | Con | 1 | 1 | 0 | 0 | 1 | 0 | 1 | 0 | 0 |
| Oliver Dowden | Con | 2 | 0 | 0 | 0 | 0 | 0 | 0 | 0 | 0 |
| Roger Gale | Con | 1 | 0 | 0 | 0 | 0 | 0 | 0 | 0 | 0 |
| Siân Berry | Green | 2 | 0 | 0 | 0 | 0 | 0 | 0 | 0 | 0 |
| Sorcha Eastwood | | 1 | 0 | 0 | 1 | 0 | 0 | 0 | 0 | 0 |
| Speaker name |  | 1 | 0 | 0 | 0 | 0 | 0 | 0 | 0 | 0 |
| Steve Witherden | | 1 | 0 | 0 | 0 | 0 | 0 | 0 | 0 | 0 |
| Tim Farron | LD | 3 | 0 | 0 | 1 | 0 | 0 | 1 | 1 | 0 |
| Tonia Antoniazzi | Lab | 1 | 0 | 0 | 0 | 0 | 0 | 0 | 0 | 0 |
| Vikki Slade |  | 2 | 1 | 0 | 1 | 1 | 0 | 0 | 1 | 0 |
| Wendy Morton | Con | 1 | 0 | 0 | 0 | 0 | 0 | 0 | 0 | 0 |
| Wera Hobhouse | LD | 2 | 0 | 0 | 0 | 0 | 0 | 0 | 0 | 0 |
| Daniel Francis | Lab | 1 | 0 | 0 | 0 | 0 | 0 | 0 | 0 | 1 |
| Danny Kruger | Con | 15 | 0 | 0 | 0 | 0 | 0 | 0 | 0 | 1 |
| Neil Shastri-Hurst | Con | 1 | 0 | 0 | 0 | 0 | 0 | 0 | 0 | 1 |
| Simon Opher |  | 1 | 0 | 0 | 0 | 0 | 0 | 0 | 0 | 1 |
| Hon. Members |  | 1 | 0 | 0 | 0 | 0 | 0 | 0 | 0 | 1 |
| Jack Abbott | Lab | 1 | 0 | 0 | 0 | 0 | 0 | 0 | 0 | 1 |
| Jake Richards |  | 2 | 0 | 0 | 0 | 0 | 0 | 0 | 0 | 1 |
| Kim Leadbeater | Lab | 11 | 0 | 0 | 0 | 0 | 0 | 0 | 0 | 1 |
| Kit Malthouse | Con | 5 | 0 | 0 | 0 | 0 | 0 | 0 | 0 | 1 |
| Lewis Atkinson | Lab | 1 | 0 | 0 | 0 | 0 | 0 | 0 | 0 | 1 |
| Lewis Atkinson | Lab | 1 | 0 | 0 | 0 | 0 | 0 | 0 | 0 | 1 |
| Liz Saville Roberts | PC | 1 | 0 | 0 | 0 | 0 | 0 | 0 | 0 | 1 |
| Naz Shah | Lab | 10 | 0 | 0 | 0 | 0 | 0 | 0 | 1 | 1 |
| Naz Shah | Lab | 1 | 0 | 0 | 0 | 0 | 0 | 0 | 0 | 1 |
| Sean Woodcock | Lab | 1 | 0 | 0 | 0 | 0 | 0 | 0 | 0 | 1 |
| Sean Woodcock | Lab | 1 | 0 | 0 | 0 | 0 | 0 | 0 | 0 | 1 |
| Sojan Joseph |  | 1 | 0 | 0 | 0 | 0 | 0 | 0 | 1 | 1 |
| Speaker name |  | 1 | 0 | 0 | 0 | 0 | 0 | 0 | 0 | 1 |
| The Chair |  | 17 | 0 | 0 | 0 | 0 | 0 | 0 | 0 | 1 |
| Tom Gordon | LD | 1 | 0 | 0 | 0 | 0 | 0 | 0 | 0 | 1 |
| Tom Gordon | LD | 1 | 0 | 0 | 0 | 0 | 0 | 0 | 0 | 1 |
| Andrew Green |  | 1 | 0 | 0 | 0 | 0 | 0 | 0 | 0 | 2 |
| Daniel Francis | Lab | 1 | 0 | 0 | 0 | 0 | 0 | 0 | 0 | 2 |
| Danny Kruger | Con | 9 | 0 | 0 | 1 | 0 | 0 | 0 | 0 | 2 |
| Green |  | 29 | 1 | 0 | 0 | 0 | 0 | 0 | 0 | 2 |
| Marie Tidball | Lab | 1 | 0 | 0 | 0 | 0 | 0 | 0 | 0 | 2 |
| Neil Shastri-Hurst | Con | 1 | 0 | 0 | 0 | 0 | 0 | 0 | 0 | 2 |
| Opher |  | 3 | 0 | 0 | 0 | 0 | 0 | 0 | 0 | 2 |
| Shastri-Hurst | Con | 1 | 0 | 0 | 0 | 0 | 0 | 0 | 0 | 2 |
| Simon Opher |  | 1 | 0 | 0 | 0 | 0 | 0 | 0 | 0 | 2 |
| Tidball | Lab | 3 | 1 | 0 | 0 | 0 | 0 | 0 | 0 | 2 |
| Duncan Burton | | 7 | 0 | 0 | 0 | 1 | 0 | 0 | 0 | 2 |
| Glyn Berry |  | 8 | 0 | 0 | 1 | 1 | 0 | 0 | 0 | 2 |
| Jack Abbott | Lab | 2 | 0 | 0 | 0 | 0 | 0 | 0 | 0 | 2 |
| Jack Abbott | Lab | 1 | 0 | 0 | 0 | 0 | 0 | 0 | 0 | 2 |
| Jake Richards |  | 5 | 0 | 0 | 0 | 0 | 0 | 0 | 0 | 2 |
| Juliet Campbell | Lab | 2 | 0 | 0 | 0 | 0 | 0 | 0 | 0 | 2 |
| Kim Leadbeater | Lab | 9 | 0 | 0 | 0 | 0 | 0 | 0 | 1 | 2 |
| Kit Malthouse | Con | 7 | 0 | 0 | 0 | 0 | 0 | 0 | 0 | 2 |
| Lewis Atkinson | Lab | 8 | 0 | 1 | 0 | 0 | 0 | 0 | 0 | 2 |
| Liz Saville Roberts | PC | 5 | 0 | 0 | 0 | 0 | 0 | 0 | 0 | 2 |
| Mark Swindells | | 13 | 0 | 0 | 0 | 0 | 0 | 0 | 0 | 2 |
| Naz Shah | Lab | 3 | 0 | 0 | 0 | 0 | 1 | 0 | 0 | 2 |
| Naz Shah | Lab | 1 | 0 | 0 | 0 | 0 | 0 | 0 | 0 | 2 |
| Nicola Ranger |  | 1 | 0 | 0 | 0 | 0 | 0 | 0 | 0 | 2 |
| Ranger |  | 13 | 0 | 0 | 0 | 0 | 0 | 0 | 1 | 2 |
| Chris Whitty |  | 1 | 0 | 0 | 0 | 0 | 0 | 0 | 0 | 2 |
| Whitty |  | 24 | 0 | 0 | 0 | 0 | 0 | 0 | 1 | 2 |
| Rebecca Paul | Con | 4 | 0 | 0 | 1 | 0 | 0 | 0 | 0 | 2 |
| Rebecca Paul | Con | 1 | 0 | 0 | 0 | 0 | 0 | 0 | 0 | 2 |
| Sarah Olney | LD | 3 | 0 | 0 | 0 | 0 | 0 | 0 | 0 | 2 |
| Sean Woodcock | Lab | 1 | 1 | 0 | 0 | 0 | 0 | 0 | 0 | 2 |
| Sojan Joseph |  | 4 | 0 | 0 | 0 | 0 | 0 | 0 | 1 | 2 |
| The Chair |  | 16 | 0 | 0 | 0 | 0 | 0 | 0 | 0 | 2 |
| Tom Gordon | LD | 2 | 0 | 0 | 0 | 0 | 0 | 0 | 0 | 2 |
| Tom Gordon | LD | 1 | 0 | 0 | 0 | 0 | 0 | 0 | 0 | 2 |
| Alex Ruck Keene | | 12 | 0 | 0 | 1 | 0 | 0 | 0 | 0 | 3 |
| Bambos Charalambous | Lab | 1 | 0 | 0 | 0 | 0 | 0 | 0 | 0 | 3 |
| Daniel Francis | Lab | 2 | 0 | 0 | 0 | 0 | 0 | 0 | 0 | 3 |
| Daniel Francis | Lab | 1 | 0 | 0 | 0 | 0 | 0 | 0 | 0 | 3 |
| Danny Kruger | Con | 11 | 1 | 0 | 0 | 0 | 0 | 0 | 0 | 3 |
| Ahmedzai |  | 9 | 0 | 0 | 0 | 0 | 0 | 0 | 1 | 3 |
| Clarke |  | 11 | 0 | 1 | 1 | 0 | 0 | 1 | 1 | 3 |
| Cox |  | 17 | 0 | 0 | 1 | 0 | 1 | 1 | 1 | 3 |
| Kaan |  | 9 | 1 | 0 | 0 | 0 | 0 | 0 | 0 | 3 |
| Marie Tidball | Lab | 1 | 0 | 0 | 0 | 0 | 0 | 0 | 0 | 3 |
| Neil Shastri-Hurst | Con | 1 | 0 | 0 | 0 | 0 | 0 | 0 | 0 | 3 |
| Opher |  | 2 | 0 | 0 | 0 | 0 | 0 | 0 | 0 | 3 |
| Rachel Clarke |  | 1 | 0 | 0 | 0 | 0 | 0 | 0 | 0 | 3 |
| Shastri-Hurst | Con | 2 | 0 | 0 | 0 | 0 | 0 | 0 | 0 | 3 |
| Simon Opher |  | 1 | 0 | 0 | 0 | 0 | 0 | 0 | 0 | 3 |
| Spielvogel |  | 18 | 1 | 0 | 0 | 0 | 0 | 0 | 0 | 3 |
| Tidball | Lab | 10 | 0 | 0 | 0 | 0 | 0 | 0 | 0 | 3 |
| Jack Abbott | Lab | 11 | 0 | 0 | 0 | 0 | 0 | 0 | 0 | 3 |
| Jack Abbott | Lab | 1 | 0 | 0 | 0 | 0 | 0 | 0 | 0 | 3 |
| Jake Richards |  | 8 | 0 | 0 | 0 | 0 | 0 | 0 | 0 | 3 |
| James Sanderson | | 5 | 0 | 0 | 0 | 0 | 0 | 0 | 0 | 3 |
| Juliet Campbell | Lab | 1 | 0 | 0 | 0 | 0 | 0 | 0 | 0 | 3 |
| Kim Leadbeater | Lab | 9 | 0 | 0 | 0 | 0 | 0 | 0 | 0 | 3 |
| Kit Malthouse | Con | 4 | 0 | 0 | 0 | 0 | 0 | 0 | 0 | 3 |
| Lewis Atkinson | Lab | 3 | 0 | 0 | 0 | 0 | 0 | 0 | 0 | 3 |
| Lewis Atkinson | Lab | 1 | 0 | 0 | 0 | 0 | 0 | 0 | 0 | 3 |
| Liz Saville Roberts | PC | 1 | 0 | 0 | 0 | 0 | 0 | 0 | 0 | 3 |
| Naz Shah | Lab | 14 | 1 | 0 | 0 | 1 | 0 | 0 | 1 | 3 |
| Rachel Hopkins | Lab | 1 | 0 | 0 | 0 | 0 | 0 | 0 | 0 | 3 |
| Rachel Hopkins | Lab | 1 | 0 | 0 | 0 | 0 | 0 | 0 | 0 | 3 |
| Rebecca Paul | Con | 2 | 0 | 0 | 0 | 0 | 0 | 0 | 0 | 3 |
| Rebecca Paul | Con | 1 | 0 | 0 | 0 | 0 | 0 | 0 | 0 | 3 |
| Sarah Cox |  | 1 | 0 | 0 | 0 | 0 | 0 | 0 | 0 | 3 |
| Sarah Green | LD | 2 | 0 | 0 | 0 | 0 | 0 | 0 | 0 | 3 |
| Sarah Olney | LD | 4 | 0 | 0 | 0 | 0 | 0 | 0 | 0 | 3 |
| Sarah Sackman | | 3 | 0 | 0 | 0 | 0 | 0 | 0 | 0 | 3 |
| Sean Woodcock | Lab | 2 | 1 | 0 | 1 | 1 | 1 | 1 | 0 | 3 |
| Max Hill |  | 16 | 1 | 0 | 0 | 0 | 0 | 0 | 0 | 3 |
| Nicholas Mostyn | | 25 | 1 | 0 | 1 | 0 | 0 | 0 | 0 | 3 |
| Sojan Joseph |  | 1 | 0 | 0 | 0 | 0 | 0 | 0 | 0 | 3 |
| Speaker name |  | 1 | 0 | 0 | 0 | 0 | 0 | 0 | 0 | 3 |
| Stephen Kinnock | Lab | 2 | 0 | 0 | 0 | 0 | 0 | 0 | 0 | 3 |
| The Chair |  | 25 | 0 | 0 | 0 | 0 | 0 | 0 | 0 | 3 |
| Tom Gordon | LD | 3 | 0 | 0 | 0 | 0 | 0 | 1 | 0 | 3 |
| Tom Gordon | LD | 1 | 0 | 0 | 0 | 0 | 0 | 0 | 0 | 3 |
| Chelsea Roff |  | 10 | 1 | 1 | 1 | 0 | 0 | 0 | 1 | 4 |
| Daniel Francis | Lab | 2 | 0 | 0 | 0 | 0 | 0 | 0 | 0 | 4 |
| Danny Kruger | Con | 3 | 0 | 0 | 0 | 0 | 0 | 0 | 0 | 4 |
| Fellingham |  | 12 | 0 | 0 | 0 | 0 | 0 | 0 | 0 | 4 |
| Griffiths |  | 8 | 0 | 1 | 0 | 0 | 0 | 0 | 0 | 4 |
| Marie Tidball | Lab | 1 | 0 | 0 | 0 | 0 | 0 | 0 | 0 | 4 |
| McLaren |  | 12 | 0 | 0 | 0 | 1 | 1 | 1 | 0 | 4 |
| Mewett |  | 13 | 0 | 0 | 0 | 0 | 0 | 0 | 0 | 4 |
| Miro Griffiths |  | 1 | 0 | 0 | 0 | 0 | 0 | 0 | 0 | 4 |
| Neil Shastri-Hurst | Con | 1 | 0 | 0 | 0 | 0 | 0 | 0 | 0 | 4 |
| Opher |  | 2 | 0 | 0 | 0 | 0 | 0 | 0 | 0 | 4 |
| Simon Opher |  | 1 | 0 | 0 | 0 | 0 | 0 | 0 | 0 | 4 |
| Tidball | Lab | 3 | 0 | 0 | 0 | 0 | 0 | 0 | 0 | 4 |
| Jack Abbott | Lab | 1 | 0 | 0 | 0 | 0 | 0 | 0 | 0 | 4 |
| Jack Abbott | Lab | 1 | 0 | 0 | 0 | 0 | 0 | 0 | 0 | 4 |
| Jake Richards |  | 2 | 0 | 0 | 0 | 0 | 0 | 0 | 0 | 4 |
| Juliet Campbell | Lab | 1 | 0 | 0 | 0 | 0 | 0 | 0 | 0 | 4 |
| Kim Leadbeater | Lab | 4 | 0 | 0 | 0 | 0 | 0 | 0 | 0 | 4 |
| Kit Malthouse | Con | 2 | 0 | 0 | 0 | 0 | 0 | 0 | 0 | 4 |
| Lewis Atkinson | Lab | 2 | 0 | 0 | 0 | 0 | 0 | 0 | 0 | 4 |
| Liz Saville Roberts | PC | 4 | 0 | 0 | 0 | 0 | 0 | 0 | 0 | 4 |
| Marie Tidball | Lab | 1 | 0 | 0 | 0 | 0 | 0 | 0 | 0 | 4 |
| Naz Shah | Lab | 4 | 0 | 0 | 1 | 0 | 0 | 0 | 1 | 4 |
| Naz Shah | Lab | 1 | 0 | 0 | 0 | 0 | 0 | 0 | 0 | 4 |
| Shakespeare |  | 7 | 0 | 0 | 0 | 0 | 0 | 0 | 1 | 4 |
| Rachel Hopkins | Lab | 1 | 0 | 0 | 0 | 0 | 0 | 0 | 0 | 4 |
| Rachel Hopkins | Lab | 1 | 0 | 0 | 0 | 0 | 0 | 0 | 1 | 4 |
| Rebecca Paul | Con | 1 | 0 | 0 | 0 | 0 | 0 | 0 | 0 | 4 |
| Sarah Green | LD | 1 | 0 | 0 | 0 | 0 | 0 | 0 | 0 | 4 |
| Sarah Olney | LD | 1 | 0 | 0 | 0 | 0 | 0 | 0 | 0 | 4 |
| Sean Woodcock | Lab | 1 | 0 | 0 | 0 | 0 | 0 | 0 | 0 | 4 |
| Simon Opher |  | 1 | 0 | 0 | 0 | 0 | 0 | 0 | 0 | 4 |
| Sojan Joseph |  | 1 | 0 | 0 | 0 | 0 | 0 | 0 | 0 | 4 |
| Speaker name |  | 1 | 0 | 0 | 0 | 0 | 0 | 0 | 0 | 4 |
| Stephen Kinnock | Lab | 2 | 0 | 0 | 0 | 0 | 0 | 0 | 0 | 4 |
| The Chair |  | 31 | 0 | 0 | 0 | 0 | 0 | 0 | 0 | 4 |
| Tom Gordon | LD | 1 | 0 | 0 | 0 | 0 | 0 | 0 | 0 | 4 |
| Tom Gordon | LD | 1 | 0 | 0 | 0 | 0 | 0 | 0 | 0 | 4 |
| Tom Shakespeare | | 1 | 0 | 0 | 0 | 0 | 0 | 0 | 0 | 4 |
| Yogi Amin |  | 12 | 0 | 0 | 0 | 0 | 0 | 0 | 0 | 4 |
| Allan House |  | 1 | 0 | 0 | 0 | 0 | 0 | 0 | 0 | 5 |
| Bambos Charalambous | Lab | 2 | 0 | 0 | 0 | 0 | 1 | 0 | 0 | 5 |
| Baroness Falkner | | 12 | 0 | 0 | 1 | 0 | 1 | 0 | 1 | 5 |
| Daniel Francis | Lab | 2 | 0 | 0 | 0 | 0 | 0 | 0 | 0 | 5 |
| Danny Kruger | Con | 6 | 0 | 0 | 0 | 0 | 0 | 0 | 1 | 5 |
| Graham |  | 7 | 0 | 0 | 0 | 0 | 0 | 0 | 0 | 5 |
| Hussain |  | 11 | 0 | 0 | 1 | 1 | 1 | 1 | 1 | 5 |
| Lewis Graham |  | 1 | 0 | 0 | 0 | 0 | 0 | 0 | 0 | 5 |
| Marie Tidball | Lab | 1 | 0 | 0 | 0 | 0 | 0 | 0 | 0 | 5 |
| Mullock |  | 10 | 1 | 0 | 1 | 0 | 0 | 1 | 0 | 5 |
| Neerkin |  | 7 | 0 | 0 | 0 | 0 | 0 | 0 | 0 | 5 |
| Opher |  | 2 | 0 | 0 | 0 | 0 | 0 | 0 | 0 | 5 |
| Simon Opher |  | 1 | 0 | 0 | 0 | 0 | 0 | 0 | 0 | 5 |
| Tidball | Lab | 4 | 0 | 0 | 0 | 0 | 0 | 0 | 0 | 5 |
| Fazilet Hadi |  | 11 | 1 | 1 | 1 | 1 | 0 | 0 | 1 | 5 |
| Jack Abbott | Lab | 2 | 0 | 0 | 0 | 0 | 0 | 0 | 0 | 5 |
| Jack Abbott | Lab | 1 | 0 | 0 | 0 | 0 | 0 | 0 | 0 | 5 |
| Jake Richards |  | 4 | 0 | 0 | 0 | 0 | 0 | 0 | 0 | 5 |
| Kim Leadbeater | Lab | 11 | 0 | 0 | 0 | 0 | 1 | 0 | 0 | 5 |
| Kit Malthouse | Con | 6 | 0 | 0 | 0 | 0 | 0 | 0 | 0 | 5 |
| Lewis Atkinson | Lab | 8 | 0 | 0 | 0 | 1 | 0 | 0 | 0 | 5 |
| Liz Saville Roberts | PC | 1 | 0 | 0 | 0 | 0 | 0 | 0 | 0 | 5 |
| Lord Sumption | | 10 | 0 | 1 | 0 | 0 | 0 | 0 | 1 | 5 |
| Naz Shah | Lab | 7 | 0 | 1 | 1 | 1 | 1 | 0 | 1 | 5 |
| Esmail |  | 7 | 0 | 0 | 0 | 0 | 0 | 0 | 0 | 5 |
| House |  | 14 | 0 | 1 | 1 | 1 | 0 | 1 | 1 | 5 |
| Rachel Hopkins | Lab | 1 | 0 | 0 | 0 | 0 | 0 | 0 | 0 | 5 |
| Rebecca Paul | Con | 4 | 0 | 0 | 0 | 0 | 0 | 0 | 0 | 5 |
| Rebecca Paul | Con | 1 | 0 | 0 | 0 | 0 | 0 | 0 | 0 | 5 |
| Richard Robinson | | 6 | 1 | 0 | 0 | 1 | 0 | 1 | 0 | 5 |
| Sam Royston |  | 7 | 0 | 0 | 1 | 1 | 0 | 0 | 0 | 5 |
| Sarah Olney | LD | 4 | 0 | 0 | 0 | 0 | 0 | 0 | 0 | 5 |
| Sarah Sackman | | 3 | 0 | 0 | 0 | 0 | 0 | 0 | 0 | 5 |
| Sean Woodcock | Lab | 5 | 0 | 0 | 1 | 1 | 1 | 0 | 0 | 5 |
| Sojan Joseph |  | 2 | 0 | 0 | 0 | 0 | 1 | 0 | 0 | 5 |
| Speaker name |  | 1 | 0 | 0 | 0 | 0 | 0 | 0 | 0 | 5 |
| Stephen Kinnock | Lab | 1 | 0 | 0 | 0 | 0 | 0 | 0 | 0 | 5 |
| The Chair |  | 22 | 0 | 0 | 0 | 0 | 0 | 0 | 0 | 5 |
| Toby Porter |  | 7 | 1 | 1 | 0 | 0 | 0 | 0 | 0 | 5 |
| Tom Gordon | LD | 2 | 0 | 0 | 0 | 0 | 0 | 0 | 0 | 5 |
| Tom Gordon | LD | 1 | 0 | 0 | 0 | 0 | 0 | 0 | 0 | 5 |
| Alex Greenwich | Ind | 11 | 0 | 0 | 0 | 0 | 0 | 0 | 1 | 6 |
| Daniel Francis | Lab | 1 | 0 | 0 | 0 | 0 | 0 | 0 | 0 | 6 |
| Danny Kruger | Con | 2 | 0 | 0 | 0 | 0 | 0 | 0 | 0 | 6 |
| Furst |  | 13 | 1 | 1 | 1 | 0 | 0 | 1 | 1 | 6 |
| Marie Tidball | Lab | 1 | 0 | 0 | 0 | 0 | 0 | 0 | 0 | 6 |
| Neil Shastri-Hurst | Con | 1 | 0 | 0 | 0 | 0 | 0 | 0 | 0 | 6 |
| Opher |  | 1 | 0 | 0 | 0 | 0 | 0 | 0 | 0 | 6 |
| Simon Opher |  | 1 | 0 | 0 | 0 | 0 | 0 | 0 | 0 | 6 |
| Jack Abbott | Lab | 1 | 0 | 0 | 0 | 0 | 0 | 0 | 0 | 6 |
| Jake Richards |  | 1 | 0 | 0 | 0 | 0 | 0 | 0 | 0 | 6 |
| Kim Leadbeater | Lab | 1 | 0 | 0 | 0 | 0 | 0 | 0 | 0 | 6 |
| Kit Malthouse | Con | 2 | 0 | 0 | 0 | 0 | 0 | 0 | 0 | 6 |
| Lewis Atkinson | Lab | 1 | 0 | 0 | 0 | 0 | 0 | 0 | 0 | 6 |
| Naz Shah | Lab | 2 | 0 | 0 | 0 | 0 | 0 | 0 | 0 | 6 |
| Blake |  | 10 | 0 | 0 | 0 | 0 | 0 | 0 | 0 | 6 |
| Rachel Hopkins | Lab | 1 | 0 | 0 | 0 | 0 | 0 | 0 | 0 | 6 |
| Sarah Green | LD | 1 | 0 | 0 | 0 | 0 | 0 | 0 | 0 | 6 |
| Sean Woodcock | Lab | 2 | 0 | 0 | 0 | 0 | 0 | 0 | 0 | 6 |
| Sean Woodcock | Lab | 1 | 0 | 0 | 0 | 0 | 0 | 0 | 0 | 6 |
| Sojan Joseph |  | 1 | 0 | 0 | 0 | 0 | 0 | 0 | 0 | 6 |
| Speaker name |  | 1 | 0 | 0 | 0 | 0 | 0 | 0 | 0 | 6 |
| The Chair |  | 9 | 0 | 0 | 0 | 0 | 0 | 0 | 0 | 6 |
| Tom Gordon | LD | 1 | 0 | 0 | 0 | 0 | 0 | 0 | 0 | 6 |
| Bambos Charalambous | Lab | 1 | 0 | 0 | 0 | 0 | 0 | 0 | 0 | 7 |
| Claire Williams | | 6 | 0 | 0 | 0 | 0 | 0 | 0 | 0 | 7 |
| Dan Scorer |  | 5 | 1 | 1 | 0 | 0 | 0 | 0 | 1 | 7 |
| Daniel Francis | Lab | 6 | 0 | 0 | 0 | 0 | 0 | 0 | 0 | 7 |
| Danny Kruger | Con | 7 | 0 | 0 | 0 | 0 | 0 | 0 | 0 | 7 |
| Amanda Ward |  | 1 | 0 | 0 | 0 | 0 | 0 | 0 | 0 | 7 |
| Annabel Price |  | 1 | 0 | 0 | 0 | 0 | 0 | 0 | 0 | 7 |
| Marie Tidball | Lab | 1 | 0 | 0 | 0 | 0 | 0 | 0 | 0 | 7 |
| Mulholland |  | 10 | 0 | 0 | 0 | 0 | 0 | 0 | 1 | 7 |
| Naomi Richards | | 1 | 0 | 0 | 0 | 0 | 0 | 0 | 0 | 7 |
| Neil Shastri-Hurst | Con | 1 | 0 | 0 | 0 | 0 | 0 | 0 | 0 | 7 |
| Opher |  | 11 | 0 | 0 | 0 | 1 | 0 | 0 | 0 | 7 |
| Price |  | 11 | 0 | 0 | 0 | 0 | 0 | 0 | 1 | 7 |
| Richards |  | 13 | 0 | 0 | 0 | 0 | 0 | 0 | 0 | 7 |
| Shastri-Hurst | Con | 2 | 0 | 0 | 0 | 0 | 0 | 0 | 0 | 7 |
| Simon Opher |  | 1 | 0 | 0 | 0 | 0 | 0 | 0 | 0 | 7 |
| Tidball | Lab | 5 | 0 | 0 | 0 | 0 | 0 | 0 | 0 | 7 |
| Ward |  | 9 | 1 | 0 | 0 | 0 | 0 | 0 | 0 | 7 |
| Emyr Lewis |  | 1 | 0 | 0 | 0 | 0 | 0 | 0 | 0 | 7 |
| Gareth Owen |  | 1 | 0 | 0 | 0 | 0 | 0 | 0 | 0 | 7 |
| Jack Abbott | Lab | 2 | 0 | 0 | 0 | 0 | 0 | 0 | 1 | 7 |
| Jack Abbott | Lab | 1 | 0 | 0 | 0 | 0 | 0 | 0 | 0 | 7 |
| Julie Thienpont | | 8 | 0 | 0 | 0 | 0 | 0 | 1 | 0 | 7 |
| Juliet Campbell | Lab | 1 | 0 | 0 | 0 | 0 | 0 | 0 | 0 | 7 |
| Kim Leadbeater | Lab | 12 | 0 | 0 | 0 | 0 | 0 | 0 | 0 | 7 |
| Kit Malthouse | Con | 6 | 0 | 0 | 0 | 0 | 0 | 0 | 0 | 7 |
| Laura Hoyano |  | 1 | 0 | 0 | 0 | 0 | 0 | 0 | 0 | 7 |
| Lewis Atkinson | Lab | 8 | 0 | 0 | 0 | 0 | 0 | 0 | 0 | 7 |
| Liz Reed |  | 10 | 0 | 0 | 0 | 0 | 0 | 0 | 0 | 7 |
| Liz Saville Roberts | PC | 3 | 0 | 0 | 0 | 0 | 0 | 0 | 0 | 7 |
| Nancy Preston |  | 1 | 0 | 0 | 0 | 0 | 0 | 0 | 0 | 7 |
| Naz Shah | Lab | 12 | 0 | 1 | 1 | 0 | 1 | 0 | 1 | 7 |
| Pat Malone |  | 13 | 1 | 0 | 1 | 0 | 0 | 1 | 1 | 7 |
| Hoyano |  | 6 | 1 | 1 | 0 | 0 | 0 | 0 | 0 | 7 |
| Hoyao |  | 1 | 0 | 0 | 0 | 0 | 0 | 0 | 0 | 7 |
| Lewis |  | 5 | 0 | 0 | 0 | 0 | 0 | 0 | 0 | 7 |
| Owen |  | 8 | 1 | 0 | 0 | 0 | 0 | 0 | 1 | 7 |
| Preston |  | 12 | 0 | 1 | 1 | 0 | 1 | 0 | 1 | 7 |
| Rachel Hopkins | Lab | 2 | 0 | 0 | 0 | 0 | 0 | 0 | 0 | 7 |
| Rachel Hopkins | Lab | 1 | 0 | 0 | 0 | 0 | 0 | 0 | 0 | 7 |
| Sarah Green | LD | 1 | 0 | 0 | 0 | 0 | 0 | 0 | 0 | 7 |
| Sarah Green | LD | 1 | 0 | 0 | 0 | 0 | 0 | 0 | 0 | 7 |
| Sarah Olney | LD | 3 | 0 | 0 | 0 | 0 | 0 | 0 | 1 | 7 |
| Sarah Olney | LD | 1 | 0 | 0 | 0 | 0 | 0 | 0 | 0 | 7 |
| Sarah Sackman | | 2 | 0 | 0 | 0 | 0 | 0 | 0 | 0 | 7 |
| Sean Woodcock | Lab | 1 | 0 | 0 | 0 | 0 | 0 | 0 | 0 | 7 |
| Simon Opher |  | 1 | 0 | 0 | 0 | 0 | 0 | 0 | 0 | 7 |
| Sojan Joseph |  | 3 | 0 | 0 | 0 | 0 | 0 | 0 | 1 | 7 |
| Speaker name |  | 1 | 0 | 0 | 0 | 0 | 0 | 0 | 0 | 7 |
| The Chair |  | 26 | 0 | 0 | 0 | 0 | 0 | 0 | 0 | 7 |
| Tom Gordon | LD | 4 | 0 | 0 | 0 | 0 | 0 | 0 | 0 | 7 |
| Tom Gordon | LD | 1 | 0 | 0 | 0 | 0 | 0 | 0 | 0 | 7 |
| Bambos Charalambous | Lab | 1 | 0 | 0 | 0 | 0 | 0 | 0 | 0 | 8 |
| Daniel Francis | Lab | 6 | 0 | 0 | 0 | 0 | 0 | 0 | 1 | 8 |
| Daniel Francis | Lab | 1 | 0 | 0 | 0 | 0 | 0 | 0 | 0 | 8 |
| Danny Kruger | Con | 17 | 1 | 0 | 0 | 0 | 0 | 0 | 1 | 8 |
| Marie Tidball | Lab | 1 | 0 | 0 | 0 | 0 | 0 | 0 | 1 | 8 |
| Neil Shastri-Hurst | Con | 1 | 0 | 0 | 0 | 0 | 0 | 0 | 0 | 8 |
| Opher |  | 4 | 0 | 0 | 0 | 0 | 0 | 0 | 1 | 8 |
| Shastri-Hurst | Con | 16 | 0 | 0 | 0 | 0 | 0 | 0 | 1 | 8 |
| Simon Opher |  | 1 | 0 | 0 | 0 | 0 | 0 | 0 | 0 | 8 |
| Tidball | Lab | 8 | 0 | 0 | 0 | 0 | 0 | 0 | 1 | 8 |
| Jack Abbott | Lab | 2 | 0 | 0 | 0 | 0 | 0 | 0 | 1 | 8 |
| Jack Abbott | Lab | 1 | 0 | 0 | 0 | 0 | 0 | 0 | 0 | 8 |
| Jake Richards |  | 3 | 0 | 0 | 0 | 0 | 0 | 0 | 1 | 8 |
| Kim Leadbeater | Lab | 14 | 0 | 0 | 0 | 0 | 0 | 0 | 0 | 8 |
| Kit Malthouse | Con | 2 | 0 | 0 | 0 | 0 | 0 | 0 | 0 | 8 |
| Lewis Atkinson | Lab | 2 | 0 | 0 | 0 | 0 | 0 | 0 | 0 | 8 |
| Liz Saville Roberts | PC | 1 | 0 | 0 | 0 | 0 | 0 | 0 | 0 | 8 |
| Naz Shah | Lab | 4 | 0 | 1 | 0 | 0 | 0 | 0 | 1 | 8 |
| Naz Shah | Lab | 1 | 0 | 0 | 0 | 0 | 0 | 0 | 0 | 8 |
| Rebecca Paul | Con | 4 | 0 | 0 | 0 | 0 | 0 | 0 | 0 | 8 |
| Sarah Olney | LD | 23 | 0 | 0 | 0 | 0 | 0 | 0 | 1 | 8 |
| Sarah Olney | LD | 1 | 0 | 0 | 0 | 0 | 0 | 0 | 0 | 8 |
| Sean Woodcock | Lab | 3 | 0 | 0 | 0 | 0 | 0 | 0 | 0 | 8 |
| Sojan Joseph |  | 2 | 0 | 0 | 0 | 0 | 0 | 0 | 0 | 8 |
| Speaker name |  | 1 | 0 | 0 | 0 | 0 | 0 | 0 | 0 | 8 |
| Stephen Kinnock | Lab | 2 | 0 | 0 | 0 | 0 | 0 | 0 | 0 | 8 |
| The Chair |  | 17 | 0 | 0 | 0 | 0 | 0 | 0 | 0 | 8 |
| Tom Gordon | LD | 1 | 0 | 0 | 0 | 0 | 0 | 0 | 0 | 8 |
| Daniel Francis | Lab | 2 | 0 | 0 | 0 | 0 | 0 | 0 | 1 | 9 |
| Danny Kruger | Con | 29 | 1 | 1 | 0 | 0 | 0 | 1 | 1 | 9 |
| Marie Tidball | Lab | 1 | 0 | 0 | 0 | 0 | 0 | 0 | 0 | 9 |
| Neil Shastri-Hurst | Con | 1 | 0 | 0 | 0 | 0 | 0 | 0 | 0 | 9 |
| Opher |  | 2 | 0 | 0 | 1 | 0 | 0 | 0 | 0 | 9 |
| Shastri-Hurst | Con | 2 | 0 | 0 | 0 | 0 | 0 | 0 | 0 | 9 |
| Simon Opher |  | 1 | 0 | 0 | 0 | 0 | 0 | 0 | 0 | 9 |
| Tidball | Lab | 1 | 0 | 0 | 0 | 0 | 0 | 0 | 1 | 9 |
| Jack Abbott | Lab | 3 | 0 | 0 | 0 | 0 | 0 | 0 | 1 | 9 |
| Jack Abbott | Lab | 1 | 0 | 0 | 0 | 0 | 0 | 0 | 0 | 9 |
| Jake Richards |  | 1 | 0 | 0 | 0 | 0 | 0 | 0 | 0 | 9 |
| Juliet Campbell | Lab | 8 | 0 | 0 | 0 | 0 | 0 | 0 | 1 | 9 |
| Kim Leadbeater | Lab | 16 | 0 | 1 | 0 | 0 | 0 | 1 | 1 | 9 |
| Kit Malthouse | Con | 18 | 0 | 0 | 0 | 0 | 0 | 0 | 1 | 9 |
| Liz Saville Roberts | PC | 1 | 0 | 0 | 0 | 0 | 0 | 0 | 0 | 9 |
| Naz Shah | Lab | 39 | 1 | 1 | 1 | 0 | 0 | 1 | 1 | 9 |
| Naz Shah | Lab | 1 | 0 | 0 | 0 | 0 | 0 | 0 | 1 | 9 |
| Rachel Hopkins | Lab | 1 | 0 | 0 | 0 | 0 | 0 | 1 | 0 | 9 |
| Rebecca Paul | Con | 2 | 0 | 0 | 1 | 1 | 0 | 0 | 0 | 9 |
| Rebecca Paul | Con | 1 | 0 | 0 | 0 | 0 | 0 | 0 | 0 | 9 |
| Sarah Olney | LD | 2 | 0 | 0 | 0 | 0 | 0 | 0 | 1 | 9 |
| Sarah Olney | LD | 1 | 0 | 0 | 0 | 0 | 0 | 0 | 0 | 9 |
| Sean Woodcock | Lab | 3 | 0 | 0 | 0 | 0 | 0 | 0 | 1 | 9 |
| Sojan Joseph |  | 4 | 0 | 1 | 1 | 0 | 0 | 0 | 1 | 9 |
| Speaker name |  | 1 | 0 | 0 | 0 | 0 | 0 | 0 | 0 | 9 |
| Stephen Kinnock | Lab | 12 | 0 | 0 | 0 | 0 | 0 | 0 | 0 | 9 |
| The Chair |  | 14 | 0 | 0 | 0 | 0 | 0 | 0 | 0 | 9 |
| Tom Gordon | LD | 3 | 0 | 0 | 0 | 0 | 0 | 0 | 0 | 9 |
| Tom Gordon | LD | 1 | 0 | 0 | 0 | 0 | 0 | 0 | 0 | 9 |
|  |  | 3 | 0 | 0 | 0 | 0 | 0 | 0 | 0 | 9 |
| Daniel Francis | Lab | 2 | 0 | 0 | 0 | 0 | 0 | 0 | 0 | 10 |
| Danny Kruger | Con | 15 | 1 | 0 | 0 | 0 | 0 | 0 | 0 | 10 |
| Marie Tidball | Lab | 1 | 0 | 0 | 0 | 0 | 0 | 0 | 0 | 10 |
| Neil Shastri-Hurst | Con | 1 | 0 | 0 | 0 | 0 | 0 | 0 | 0 | 10 |
| Opher |  | 1 | 0 | 0 | 0 | 0 | 0 | 0 | 0 | 10 |
| Simon Opher |  | 1 | 0 | 0 | 0 | 0 | 0 | 0 | 0 | 10 |
| Tidball | Lab | 2 | 0 | 0 | 0 | 0 | 0 | 0 | 0 | 10 |
| Jake Richards |  | 11 | 1 | 0 | 0 | 0 | 0 | 1 | 0 | 10 |
| Juliet Campbell | Lab | 5 | 0 | 0 | 0 | 0 | 0 | 0 | 0 | 10 |
| Kim Leadbeater | Lab | 10 | 1 | 0 | 0 | 0 | 0 | 0 | 0 | 10 |
| Kit Malthouse | Con | 3 | 1 | 0 | 0 | 0 | 0 | 0 | 0 | 10 |
| Lewis Atkinson | Lab | 1 | 0 | 0 | 0 | 0 | 0 | 0 | 0 | 10 |
| Liz Saville Roberts | PC | 1 | 0 | 0 | 0 | 0 | 0 | 0 | 0 | 10 |
| Naz Shah | Lab | 18 | 1 | 1 | 0 | 0 | 0 | 0 | 1 | 10 |
| Naz Shah | Lab | 1 | 0 | 0 | 0 | 0 | 0 | 0 | 0 | 10 |
| Rachel Hopkins | Lab | 2 | 0 | 0 | 0 | 0 | 0 | 0 | 0 | 10 |
| Rachel Hopkins | Lab | 1 | 0 | 0 | 0 | 0 | 0 | 0 | 0 | 10 |
| Rebecca Paul | Con | 36 | 1 | 0 | 0 | 0 | 0 | 0 | 1 | 10 |
| Rebecca Paul | Con | 1 | 0 | 0 | 0 | 0 | 0 | 0 | 0 | 10 |
| Sarah Olney | LD | 4 | 1 | 0 | 0 | 0 | 0 | 0 | 0 | 10 |
| Sarah Sackman | | 1 | 0 | 0 | 0 | 0 | 0 | 0 | 0 | 10 |
| Sean Woodcock | Lab | 3 | 1 | 0 | 0 | 0 | 0 | 0 | 0 | 10 |
| Sojan Joseph |  | 1 | 0 | 1 | 1 | 0 | 0 | 0 | 0 | 10 |
| Speaker name |  | 1 | 0 | 0 | 0 | 0 | 0 | 0 | 0 | 10 |
| Stephen Kinnock | Lab | 1 | 0 | 0 | 0 | 0 | 0 | 0 | 0 | 10 |
| The Chair |  | 4 | 0 | 0 | 0 | 0 | 0 | 0 | 0 | 10 |
| Tom Gordon | LD | 1 | 0 | 0 | 0 | 0 | 0 | 0 | 0 | 10 |
| Bambos Charalambous | Lab | 1 | 0 | 0 | 0 | 0 | 0 | 0 | 0 | 11 |
| Daniel Francis | Lab | 2 | 1 | 0 | 0 | 0 | 0 | 0 | 0 | 11 |
| Danny Kruger | Con | 42 | 1 | 0 | 1 | 1 | 0 | 1 | 0 | 11 |
| Danny Kruger | Con | 1 | 0 | 0 | 0 | 0 | 0 | 0 | 0 | 11 |
| Aneez Esmail |  | 1 | 0 | 0 | 0 | 0 | 0 | 0 | 0 | 11 |
| Marie Tidball | Lab | 1 | 0 | 0 | 0 | 0 | 0 | 0 | 0 | 11 |
| Neil Shastri-Hurst | Con | 1 | 0 | 0 | 0 | 0 | 0 | 0 | 0 | 11 |
| Opher |  | 7 | 0 | 0 | 0 | 1 | 0 | 1 | 0 | 11 |
| Shastri-Hurst | Con | 3 | 0 | 0 | 0 | 0 | 0 | 0 | 0 | 11 |
| Simon Opher |  | 1 | 0 | 0 | 0 | 0 | 0 | 0 | 0 | 11 |
| Tidball | Lab | 3 | 0 | 0 | 0 | 0 | 0 | 0 | 0 | 11 |
| Jack Abbott | Lab | 1 | 0 | 0 | 0 | 0 | 0 | 0 | 0 | 11 |
| Jack Abbott | Lab | 1 | 0 | 0 | 0 | 0 | 0 | 0 | 0 | 11 |
| Jake Richards |  | 9 | 0 | 1 | 0 | 0 | 0 | 0 | 0 | 11 |
| Juliet Campbell | Lab | 1 | 0 | 0 | 0 | 0 | 0 | 0 | 0 | 11 |
| Kim Leadbeater | Lab | 17 | 1 | 0 | 0 | 0 | 0 | 1 | 1 | 11 |
| Kit Malthouse | Con | 14 | 0 | 0 | 0 | 1 | 0 | 0 | 0 | 11 |
| Liz Saville Roberts | PC | 1 | 0 | 0 | 0 | 0 | 0 | 0 | 0 | 11 |
| Naz Shah | Lab | 33 | 1 | 1 | 0 | 1 | 1 | 1 | 1 | 11 |
| Naz Shah | Lab | 1 | 1 | 0 | 0 | 0 | 0 | 0 | 0 | 11 |
| Rachel Hopkins | Lab | 2 | 0 | 0 | 0 | 0 | 0 | 0 | 0 | 11 |
| Rachel Hopkins | Lab | 1 | 0 | 0 | 0 | 0 | 0 | 0 | 0 | 11 |
| Rebecca Paul | Con | 9 | 0 | 0 | 0 | 1 | 0 | 0 | 0 | 11 |
| Rebecca Paul | Con | 1 | 0 | 0 | 0 | 0 | 0 | 0 | 0 | 11 |
| Sarah Olney | LD | 2 | 0 | 0 | 0 | 1 | 0 | 0 | 0 | 11 |
| Sarah Sackman | | 5 | 1 | 0 | 0 | 0 | 0 | 0 | 0 | 11 |
| Sean Woodcock | Lab | 4 | 0 | 0 | 1 | 1 | 0 | 1 | 0 | 11 |
| Sean Woodcock | Lab | 1 | 0 | 0 | 0 | 0 | 0 | 0 | 0 | 11 |
| Nicholas Mostyn | | 1 | 0 | 0 | 0 | 0 | 0 | 0 | 0 | 11 |
| Sojan Joseph |  | 1 | 0 | 0 | 0 | 0 | 0 | 0 | 1 | 11 |
| Speaker name |  | 2 | 0 | 0 | 0 | 0 | 0 | 0 | 0 | 11 |
| Stephen Kinnock | Lab | 3 | 0 | 0 | 0 | 0 | 0 | 0 | 0 | 11 |
| The Chair |  | 16 | 0 | 0 | 0 | 0 | 0 | 0 | 0 | 11 |
| Tom Gordon | LD | 3 | 0 | 0 | 0 | 0 | 0 | 0 | 0 | 11 |
| Tom Gordon | LD | 1 | 0 | 0 | 0 | 0 | 0 | 0 | 0 | 11 |
| Daniel Francis | Lab | 1 | 0 | 0 | 0 | 0 | 1 | 0 | 0 | 12 |
| Danny Kruger | Con | 20 | 0 | 0 | 1 | 1 | 0 | 1 | 1 | 12 |
| Neil Shastri-Hurst | Con | 1 | 0 | 0 | 0 | 0 | 0 | 0 | 0 | 12 |
| Opher |  | 2 | 0 | 0 | 0 | 0 | 0 | 0 | 0 | 12 |
| Shastri-Hurst | Con | 10 | 0 | 0 | 0 | 0 | 0 | 0 | 0 | 12 |
| Simon Opher |  | 1 | 0 | 0 | 0 | 0 | 0 | 0 | 0 | 12 |
| Jack Abbott | Lab | 1 | 0 | 0 | 0 | 0 | 0 | 0 | 0 | 12 |
| Jack Abbott | Lab | 1 | 0 | 0 | 0 | 0 | 0 | 0 | 0 | 12 |
| Juliet Campbell | Lab | 2 | 0 | 0 | 0 | 0 | 0 | 0 | 0 | 12 |
| Kim Leadbeater | Lab | 18 | 0 | 0 | 1 | 0 | 0 | 0 | 1 | 12 |
| Kit Malthouse | Con | 3 | 0 | 0 | 0 | 0 | 0 | 0 | 0 | 12 |
| Lewis Atkinson | Lab | 1 | 0 | 0 | 0 | 0 | 0 | 0 | 0 | 12 |
| Naz Shah | Lab | 22 | 0 | 0 | 1 | 0 | 1 | 0 | 1 | 12 |
| Naz Shah | Lab | 1 | 0 | 0 | 0 | 0 | 0 | 0 | 0 | 12 |
| Rebecca Paul | Con | 9 | 0 | 1 | 1 | 1 | 0 | 0 | 1 | 12 |
| Sarah Green | LD | 1 | 0 | 0 | 0 | 0 | 0 | 0 | 1 | 12 |
| Sarah Olney | LD | 1 | 0 | 0 | 0 | 0 | 0 | 0 | 0 | 12 |
| Sean Woodcock | Lab | 1 | 0 | 0 | 0 | 0 | 0 | 0 | 0 | 12 |
| Sojan Joseph |  | 1 | 0 | 0 | 0 | 0 | 0 | 0 | 0 | 12 |
| Speaker name |  | 1 | 0 | 0 | 0 | 0 | 0 | 0 | 0 | 12 |
| Stephen Kinnock | Lab | 2 | 0 | 0 | 0 | 0 | 0 | 0 | 0 | 12 |
| The Chair |  | 11 | 0 | 0 | 0 | 0 | 0 | 0 | 0 | 12 |
| Tom Gordon | LD | 1 | 0 | 0 | 0 | 0 | 0 | 0 | 0 | 12 |
| Bambos Charalambous | Lab | 1 | 0 | 0 | 0 | 0 | 0 | 0 | 0 | 13 |
| Daniel Francis | Lab | 4 | 0 | 0 | 0 | 0 | 0 | 0 | 0 | 13 |
| Danny Kruger | Con | 15 | 0 | 1 | 0 | 0 | 0 | 0 | 1 | 13 |
| Marie Tidball | Lab | 1 | 0 | 0 | 0 | 0 | 0 | 0 | 0 | 13 |
| Neil Shastri-Hurst | Con | 1 | 0 | 0 | 0 | 0 | 0 | 0 | 0 | 13 |
| Opher |  | 6 | 0 | 0 | 0 | 0 | 0 | 0 | 0 | 13 |
| Shastri-Hurst | Con | 2 | 0 | 0 | 0 | 0 | 0 | 0 | 0 | 13 |
| Simon Opher |  | 1 | 0 | 0 | 0 | 0 | 0 | 0 | 0 | 13 |
| Tidball | Lab | 3 | 0 | 0 | 0 | 0 | 0 | 0 | 0 | 13 |
| Jack Abbott | Lab | 3 | 0 | 0 | 0 | 0 | 0 | 0 | 0 | 13 |
| Jack Abbott | Lab | 1 | 0 | 0 | 0 | 0 | 0 | 0 | 0 | 13 |
| Jake Richards |  | 1 | 0 | 0 | 0 | 0 | 0 | 0 | 0 | 13 |
| Juliet Campbell | Lab | 4 | 0 | 0 | 0 | 0 | 0 | 0 | 0 | 13 |
| Kim Leadbeater | Lab | 13 | 0 | 0 | 0 | 0 | 0 | 0 | 1 | 13 |
| Kit Malthouse | Con | 6 | 0 | 1 | 0 | 0 | 0 | 0 | 1 | 13 |
| Lewis Atkinson | Lab | 9 | 0 | 0 | 0 | 0 | 0 | 0 | 1 | 13 |
| Naz Shah | Lab | 30 | 0 | 1 | 0 | 0 | 0 | 0 | 1 | 13 |
| Naz Shah | Lab | 1 | 0 | 0 | 0 | 0 | 0 | 0 | 1 | 13 |
| Rachel Hopkins | Lab | 1 | 0 | 0 | 0 | 0 | 0 | 0 | 0 | 13 |
| Rebecca Paul | Con | 6 | 0 | 1 | 1 | 1 | 0 | 0 | 1 | 13 |
| Rebecca Paul | Con | 1 | 0 | 0 | 0 | 0 | 0 | 0 | 0 | 13 |
| Sarah Olney | LD | 2 | 0 | 0 | 0 | 0 | 0 | 0 | 0 | 13 |
| Sean Woodcock | Lab | 1 | 0 | 0 | 0 | 0 | 0 | 0 | 0 | 13 |
| Speaker name |  | 2 | 0 | 0 | 0 | 0 | 0 | 0 | 0 | 13 |
| Stephen Kinnock | Lab | 10 | 0 | 0 | 0 | 0 | 0 | 0 | 1 | 13 |
| The Chair |  | 11 | 0 | 0 | 0 | 0 | 0 | 0 | 0 | 13 |
| Tom Gordon | LD | 3 | 0 | 0 | 0 | 0 | 0 | 0 | 0 | 13 |
| Tom Gordon | LD | 1 | 0 | 0 | 0 | 0 | 0 | 0 | 0 | 13 |
| Daniel Francis | Lab | 3 | 1 | 1 | 0 | 0 | 0 | 0 | 1 | 14 |
| Daniel Francis | Lab | 1 | 0 | 0 | 0 | 0 | 0 | 0 | 0 | 14 |
| Danny Kruger | Con | 15 | 0 | 0 | 0 | 1 | 0 | 1 | 1 | 14 |
| Opher |  | 12 | 0 | 0 | 0 | 0 | 0 | 0 | 1 | 14 |
| Simon Opher |  | 1 | 0 | 0 | 0 | 0 | 0 | 0 | 1 | 14 |
| Jack Abbott | Lab | 1 | 0 | 0 | 0 | 0 | 0 | 0 | 0 | 14 |
| Jack Abbott | Lab | 1 | 0 | 0 | 0 | 0 | 0 | 0 | 0 | 14 |
| Jake Richards |  | 9 | 0 | 0 | 0 | 0 | 0 | 0 | 0 | 14 |
| Kim Leadbeater | Lab | 5 | 0 | 0 | 0 | 0 | 0 | 0 | 0 | 14 |
| Kit Malthouse | Con | 1 | 0 | 0 | 0 | 0 | 0 | 0 | 0 | 14 |
| Lewis Atkinson | Lab | 3 | 0 | 0 | 0 | 0 | 0 | 0 | 0 | 14 |
| Liz Saville Roberts | PC | 1 | 0 | 0 | 0 | 0 | 0 | 0 | 0 | 14 |
| Naz Shah | Lab | 14 | 0 | 1 | 1 | 0 | 0 | 0 | 1 | 14 |
| Rebecca Paul | Con | 1 | 0 | 1 | 0 | 0 | 0 | 0 | 1 | 14 |
| Sarah Olney | LD | 4 | 0 | 0 | 0 | 0 | 0 | 0 | 1 | 14 |
| Sojan Joseph |  | 4 | 0 | 0 | 0 | 0 | 0 | 0 | 1 | 14 |
| Speaker name |  | 1 | 0 | 0 | 0 | 0 | 0 | 0 | 0 | 14 |
| The Chair |  | 8 | 0 | 0 | 0 | 0 | 0 | 0 | 0 | 14 |
| Tom Gordon | LD | 1 | 0 | 0 | 0 | 0 | 0 | 0 | 0 | 14 |
| Daniel Francis | Lab | 5 | 1 | 1 | 0 | 0 | 0 | 0 | 1 | 15 |
| Danny Kruger | Con | 30 | 1 | 1 | 0 | 0 | 0 | 1 | 1 | 15 |
| Marie Tidball | Lab | 1 | 0 | 0 | 0 | 0 | 0 | 0 | 1 | 15 |
| Neil Shastri-Hurst | Con | 1 | 0 | 0 | 0 | 0 | 0 | 0 | 1 | 15 |
| Opher |  | 5 | 0 | 0 | 0 | 0 | 0 | 0 | 0 | 15 |
| Shastri-Hurst | Con | 2 | 0 | 0 | 0 | 0 | 0 | 0 | 0 | 15 |
| Simon Opher |  | 1 | 0 | 0 | 0 | 0 | 0 | 0 | 0 | 15 |
| Tidball | Lab | 3 | 0 | 0 | 0 | 0 | 0 | 0 | 0 | 15 |
| Jack Abbott | Lab | 5 | 0 | 0 | 0 | 0 | 0 | 0 | 0 | 15 |
| Jack Abbott | Lab | 1 | 0 | 0 | 0 | 0 | 0 | 0 | 0 | 15 |
| Jake Richards |  | 2 | 0 | 0 | 0 | 0 | 0 | 0 | 0 | 15 |
| Juliet Campbell | Lab | 2 | 0 | 1 | 1 | 0 | 1 | 0 | 1 | 15 |
| Kim Leadbeater | Lab | 11 | 0 | 0 | 0 | 0 | 0 | 0 | 0 | 15 |
| Kit Malthouse | Con | 7 | 0 | 0 | 0 | 1 | 0 | 1 | 0 | 15 |
| Lewis Atkinson | Lab | 6 | 0 | 0 | 0 | 0 | 0 | 0 | 0 | 15 |
| Marie Tidball | Lab | 1 | 0 | 0 | 0 | 0 | 0 | 0 | 0 | 15 |
| Naz Shah | Lab | 13 | 0 | 1 | 1 | 0 | 1 | 0 | 1 | 15 |
| Rebecca Paul | Con | 5 | 0 | 0 | 0 | 0 | 0 | 0 | 0 | 15 |
| Rebecca Paul | Con | 1 | 0 | 0 | 0 | 0 | 0 | 0 | 0 | 15 |
| Sarah Green | LD | 1 | 0 | 0 | 0 | 0 | 0 | 0 | 0 | 15 |
| Sarah Green | LD | 1 | 0 | 0 | 0 | 0 | 0 | 0 | 0 | 15 |
| Sarah Olney | LD | 3 | 0 | 0 | 0 | 0 | 0 | 1 | 1 | 15 |
| Sean Woodcock | Lab | 1 | 0 | 0 | 0 | 0 | 0 | 0 | 0 | 15 |
| Sojan Joseph |  | 5 | 1 | 1 | 0 | 0 | 0 | 0 | 1 | 15 |
| Speaker name |  | 2 | 0 | 0 | 0 | 0 | 0 | 0 | 0 | 15 |
| Stephen Kinnock | Lab | 16 | 0 | 0 | 0 | 0 | 0 | 0 | 0 | 15 |
| The Chair |  | 10 | 0 | 0 | 0 | 0 | 0 | 0 | 0 | 15 |
| Tom Gordon | LD | 1 | 0 | 0 | 0 | 0 | 0 | 0 | 0 | 15 |
| Daniel Francis | Lab | 2 | 0 | 0 | 0 | 0 | 0 | 0 | 0 | 16 |
| Danny Kruger | Con | 14 | 0 | 0 | 0 | 0 | 0 | 0 | 0 | 16 |
| Marie Tidball | Lab | 1 | 0 | 0 | 0 | 0 | 0 | 0 | 0 | 16 |
| Opher |  | 7 | 0 | 0 | 0 | 0 | 0 | 0 | 0 | 16 |
| Simon Opher |  | 1 | 0 | 0 | 0 | 0 | 0 | 0 | 0 | 16 |
| Tidball | Lab | 6 | 0 | 0 | 0 | 0 | 0 | 0 | 0 | 16 |
| Jake Richards |  | 1 | 0 | 0 | 0 | 0 | 0 | 0 | 0 | 16 |
| Juliet Campbell | Lab | 1 | 0 | 0 | 0 | 0 | 0 | 0 | 0 | 16 |
| Kim Leadbeater | Lab | 11 | 0 | 0 | 0 | 0 | 0 | 0 | 0 | 16 |
| Kit Malthouse | Con | 4 | 0 | 0 | 0 | 0 | 0 | 0 | 0 | 16 |
| Lewis Atkinson | Lab | 4 | 0 | 0 | 0 | 0 | 0 | 0 | 0 | 16 |
| Naz Shah | Lab | 6 | 0 | 0 | 0 | 0 | 0 | 0 | 0 | 16 |
| Naz Shah | Lab | 1 | 0 | 0 | 0 | 0 | 0 | 0 | 0 | 16 |
| Rebecca Paul | Con | 3 | 0 | 1 | 0 | 0 | 0 | 0 | 0 | 16 |
| Rebecca Paul | Con | 1 | 0 | 0 | 0 | 0 | 0 | 0 | 0 | 16 |
| Sarah Green | LD | 1 | 0 | 0 | 0 | 0 | 0 | 0 | 0 | 16 |
| Sean Woodcock | Lab | 1 | 0 | 0 | 0 | 0 | 0 | 0 | 0 | 16 |
| Sojan Joseph |  | 2 | 0 | 1 | 0 | 0 | 0 | 0 | 1 | 16 |
| Speaker name |  | 2 | 0 | 0 | 0 | 0 | 0 | 0 | 0 | 16 |
| Stephen Kinnock | Lab | 6 | 0 | 0 | 0 | 0 | 0 | 0 | 0 | 16 |
| The Chair |  | 14 | 0 | 0 | 0 | 0 | 0 | 0 | 0 | 16 |
|  |  | 1 | 0 | 0 | 0 | 0 | 0 | 0 | 0 | 16 |
| Daniel Francis | Lab | 3 | 0 | 0 | 0 | 0 | 0 | 0 | 0 | 17 |
| Danny Kruger | Con | 20 | 1 | 0 | 0 | 1 | 1 | 0 | 1 | 17 |
| Neil Shastri-Hurst | Con | 1 | 0 | 0 | 0 | 0 | 0 | 0 | 0 | 17 |
| Shastri-Hurst | Con | 1 | 0 | 0 | 0 | 0 | 0 | 0 | 0 | 17 |
| Juliet Campbell | Lab | 5 | 0 | 0 | 0 | 0 | 0 | 0 | 0 | 17 |
| Juliet Campbell | Lab | 1 | 0 | 0 | 0 | 0 | 0 | 0 | 0 | 17 |
| Kim Leadbeater | Lab | 12 | 0 | 0 | 0 | 0 | 0 | 0 | 0 | 17 |
| Kit Malthouse | Con | 11 | 0 | 0 | 0 | 0 | 0 | 0 | 0 | 17 |
| Lewis Atkinson | Lab | 5 | 0 | 0 | 0 | 0 | 0 | 0 | 0 | 17 |
| Liz Saville Roberts | PC | 6 | 0 | 0 | 0 | 0 | 0 | 0 | 0 | 17 |
| Naz Shah | Lab | 13 | 0 | 1 | 0 | 0 | 1 | 0 | 1 | 17 |
| Rebecca Paul | Con | 3 | 0 | 0 | 0 | 0 | 0 | 0 | 0 | 17 |
| Rebecca Paul | Con | 1 | 0 | 0 | 0 | 0 | 0 | 0 | 0 | 17 |
| Sarah Green | LD | 1 | 0 | 0 | 0 | 0 | 0 | 0 | 0 | 17 |
| Sarah Olney | LD | 3 | 0 | 0 | 0 | 0 | 0 | 1 | 1 | 17 |
| Sarah Olney | LD | 1 | 0 | 0 | 0 | 0 | 0 | 0 | 0 | 17 |
| Sean Woodcock | Lab | 4 | 0 | 1 | 0 | 0 | 1 | 0 | 1 | 17 |
| Sojan Joseph |  | 4 | 0 | 0 | 0 | 0 | 0 | 0 | 1 | 17 |
| Speaker name |  | 7 | 0 | 0 | 0 | 0 | 0 | 0 | 0 | 17 |
| Stephen Kinnock | Lab | 18 | 0 | 0 | 0 | 0 | 0 | 0 | 0 | 17 |
| The Chair |  | 28 | 0 | 0 | 0 | 0 | 0 | 0 | 0 | 17 |
| Daniel Francis | Lab | 3 | 0 | 0 | 0 | 0 | 0 | 0 | 0 | 18 |
| Danny Kruger | Con | 14 | 0 | 1 | 0 | 0 | 0 | 0 | 1 | 18 |
| Marie Tidball | Lab | 1 | 0 | 0 | 0 | 0 | 0 | 0 | 1 | 18 |
| Neil Shastri-Hurst | Con | 1 | 0 | 0 | 0 | 0 | 0 | 0 | 0 | 18 |
| Opher |  | 2 | 0 | 0 | 0 | 0 | 0 | 0 | 0 | 18 |
| Simon Opher |  | 1 | 0 | 0 | 0 | 0 | 0 | 0 | 0 | 18 |
| Tidball | Lab | 1 | 0 | 0 | 0 | 0 | 0 | 0 | 1 | 18 |
| Jack Abbott | Lab | 2 | 0 | 0 | 0 | 0 | 0 | 0 | 0 | 18 |
| Jack Abbott | Lab | 1 | 0 | 0 | 0 | 0 | 0 | 0 | 0 | 18 |
| Kim Leadbeater | Lab | 12 | 0 | 0 | 0 | 1 | 0 | 0 | 0 | 18 |
| Kit Malthouse | Con | 1 | 0 | 0 | 0 | 0 | 0 | 0 | 0 | 18 |
| Liz Saville Roberts | PC | 10 | 0 | 0 | 0 | 0 | 0 | 0 | 0 | 18 |
| Naz Shah | Lab | 14 | 1 | 0 | 0 | 1 | 1 | 1 | 1 | 18 |
| Rachel Hopkins | Lab | 1 | 0 | 0 | 0 | 0 | 0 | 0 | 0 | 18 |
| Rebecca Paul | Con | 1 | 0 | 0 | 0 | 0 | 0 | 0 | 0 | 18 |
| Rebecca Paul | Con | 1 | 0 | 0 | 0 | 0 | 0 | 0 | 0 | 18 |
| Speaker name |  | 3 | 0 | 0 | 0 | 0 | 0 | 0 | 0 | 18 |
| Stephen Kinnock | Lab | 13 | 0 | 0 | 0 | 0 | 0 | 0 | 0 | 18 |
| The Chair |  | 11 | 0 | 0 | 0 | 0 | 0 | 0 | 0 | 18 |
| Daniel Francis | Lab | 8 | 0 | 0 | 0 | 0 | 0 | 0 | 1 | 19 |
| Danny Kruger | Con | 36 | 0 | 1 | 0 | 0 | 0 | 0 | 1 | 19 |
| Neil Shastri-Hurst | Con | 1 | 0 | 0 | 0 | 0 | 0 | 0 | 0 | 19 |
| Opher |  | 11 | 0 | 0 | 0 | 0 | 0 | 0 | 0 | 19 |
| Simon Opher |  | 1 | 0 | 0 | 0 | 0 | 0 | 0 | 0 | 19 |
| Jack Abbott | Lab | 2 | 0 | 0 | 0 | 0 | 0 | 0 | 0 | 19 |
| Jack Abbott | Lab | 1 | 0 | 0 | 0 | 0 | 0 | 0 | 0 | 19 |
| Jake Richards |  | 8 | 0 | 0 | 0 | 0 | 0 | 0 | 1 | 19 |
| Juliet Campbell | Lab | 1 | 0 | 0 | 0 | 0 | 0 | 0 | 0 | 19 |
| Kim Leadbeater | Lab | 26 | 0 | 0 | 0 | 0 | 0 | 0 | 1 | 19 |
| Kit Malthouse | Con | 12 | 0 | 0 | 0 | 0 | 0 | 0 | 0 | 19 |
| Lewis Atkinson | Lab | 8 | 0 | 1 | 0 | 0 | 0 | 0 | 1 | 19 |
| Liz Saville Roberts | PC | 1 | 0 | 0 | 0 | 0 | 0 | 0 | 0 | 19 |
| Naz Shah | Lab | 16 | 0 | 0 | 0 | 0 | 1 | 0 | 1 | 19 |
| Rebecca Paul | Con | 8 | 0 | 0 | 0 | 0 | 0 | 0 | 1 | 19 |
| Rebecca Paul | Con | 1 | 0 | 0 | 0 | 0 | 0 | 0 | 0 | 19 |
| Sarah Olney | LD | 10 | 0 | 0 | 0 | 0 | 0 | 0 | 1 | 19 |
| Sean Woodcock | Lab | 2 | 0 | 0 | 0 | 0 | 0 | 0 | 0 | 19 |
| Sojan Joseph |  | 9 | 0 | 0 | 0 | 0 | 0 | 0 | 1 | 19 |
| Speaker name |  | 4 | 0 | 0 | 0 | 0 | 0 | 0 | 0 | 19 |
| Stephen Kinnock | Lab | 13 | 0 | 1 | 0 | 0 | 0 | 0 | 1 | 19 |
| Steve Kinnock | Lab | 1 | 0 | 0 | 0 | 0 | 0 | 0 | 0 | 19 |
| The Chair |  | 33 | 0 | 0 | 0 | 0 | 0 | 0 | 1 | 19 |
| Danny Kruger | Con | 14 | 1 | 0 | 0 | 1 | 0 | 1 | 1 | 20 |
| Danny Kruger | Con | 1 | 0 | 0 | 0 | 0 | 0 | 0 | 0 | 20 |
| Opher |  | 5 | 0 | 0 | 0 | 0 | 0 | 0 | 0 | 20 |
| Simon Opher |  | 1 | 0 | 0 | 0 | 0 | 0 | 0 | 0 | 20 |
| Jack Abbott | Lab | 3 | 0 | 0 | 0 | 0 | 0 | 0 | 0 | 20 |
| Jack Abbott | Lab | 1 | 0 | 0 | 0 | 0 | 0 | 0 | 0 | 20 |
| Jake Richards |  | 3 | 0 | 0 | 0 | 0 | 0 | 0 | 0 | 20 |
| Juliet Campbell | Lab | 1 | 0 | 0 | 0 | 0 | 0 | 0 | 0 | 20 |
| Kim Leadbeater | Lab | 6 | 0 | 0 | 0 | 1 | 0 | 0 | 0 | 20 |
| Kit Malthouse | Con | 3 | 0 | 0 | 0 | 0 | 0 | 0 | 0 | 20 |
| Lewis Atkinson | Lab | 4 | 0 | 0 | 0 | 0 | 0 | 0 | 0 | 20 |
| Naz Shah | Lab | 18 | 0 | 0 | 0 | 0 | 0 | 0 | 1 | 20 |
| Rachel Hopkins | Lab | 7 | 0 | 0 | 0 | 0 | 0 | 0 | 0 | 20 |
| Rebecca Paul | Con | 4 | 0 | 0 | 0 | 0 | 0 | 0 | 0 | 20 |
| Rebecca Paul | Con | 1 | 0 | 0 | 0 | 0 | 0 | 0 | 0 | 20 |
| Sarah Olney | LD | 3 | 0 | 0 | 0 | 0 | 0 | 0 | 0 | 20 |
| Sojan Joseph |  | 2 | 0 | 0 | 0 | 0 | 0 | 0 | 1 | 20 |
| Speaker name |  | 1 | 0 | 0 | 0 | 0 | 0 | 0 | 0 | 20 |
| Stephen Kinnock | Lab | 3 | 0 | 0 | 0 | 0 | 0 | 0 | 0 | 20 |
| The Chair |  | 6 | 0 | 0 | 0 | 0 | 0 | 0 | 0 | 20 |
| Daniel Francis | Lab | 9 | 1 | 0 | 0 | 0 | 0 | 0 | 1 | 21 |
| Danny Kruger | Con | 33 | 1 | 0 | 0 | 0 | 0 | 0 | 0 | 21 |
| Marie Tidball | Lab | 1 | 0 | 0 | 0 | 0 | 0 | 0 | 0 | 21 |
| Neil Shastri-Hurst | Con | 1 | 0 | 0 | 0 | 0 | 0 | 0 | 0 | 21 |
| Opher |  | 6 | 0 | 0 | 0 | 0 | 0 | 0 | 0 | 21 |
| Shastri-Hurst | Con | 4 | 0 | 0 | 0 | 0 | 0 | 0 | 0 | 21 |
| Simon Opher |  | 1 | 0 | 0 | 0 | 0 | 0 | 0 | 0 | 21 |
| Jack Abbott | Lab | 5 | 0 | 0 | 0 | 0 | 0 | 0 | 0 | 21 |
| Jack Abbott | Lab | 1 | 0 | 0 | 0 | 0 | 0 | 0 | 0 | 21 |
| Jake Richards |  | 17 | 0 | 0 | 0 | 0 | 0 | 0 | 0 | 21 |
| Juliet Campbell | Lab | 5 | 1 | 0 | 0 | 1 | 0 | 1 | 0 | 21 |
| Kim Leadbeater | Lab | 26 | 0 | 0 | 0 | 0 | 0 | 0 | 0 | 21 |
| Kit Malthouse | Con | 2 | 0 | 0 | 0 | 0 | 0 | 0 | 0 | 21 |
| Lewis Atkinson | Lab | 8 | 0 | 0 | 0 | 0 | 0 | 0 | 1 | 21 |
| Liz Saville Roberts | PC | 1 | 0 | 0 | 0 | 0 | 0 | 0 | 0 | 21 |
| Naz Shah | Lab | 32 | 1 | 1 | 1 | 0 | 0 | 1 | 1 | 21 |
| Rachel Hopkins | Lab | 1 | 0 | 0 | 0 | 0 | 0 | 0 | 0 | 21 |
| Rachel Hopkins | Lab | 1 | 0 | 0 | 0 | 0 | 0 | 0 | 0 | 21 |
| Rebecca Paul | Con | 8 | 1 | 0 | 0 | 0 | 0 | 0 | 0 | 21 |
| Rebecca Paul | Con | 1 | 0 | 0 | 0 | 0 | 0 | 0 | 0 | 21 |
| Sarah Green | LD | 1 | 0 | 0 | 0 | 0 | 0 | 0 | 0 | 21 |
| Sarah Green | LD | 1 | 0 | 0 | 0 | 0 | 0 | 0 | 0 | 21 |
| Sarah Olney | LD | 9 | 0 | 0 | 0 | 0 | 0 | 0 | 0 | 21 |
| Sarah Olney | LD | 1 | 0 | 0 | 0 | 0 | 0 | 0 | 0 | 21 |
| Sarah Sackman | | 4 | 0 | 0 | 0 | 0 | 0 | 0 | 0 | 21 |
| Sean Woodcock | Lab | 2 | 1 | 0 | 0 | 0 | 0 | 0 | 0 | 21 |
| Sojan Joseph |  | 2 | 0 | 0 | 0 | 0 | 0 | 0 | 0 | 21 |
| Speaker name |  | 5 | 0 | 0 | 0 | 0 | 0 | 0 | 0 | 21 |
| Stephen Kinnock | Lab | 10 | 0 | 0 | 0 | 0 | 0 | 0 | 0 | 21 |
| The Chair |  | 17 | 0 | 0 | 0 | 0 | 0 | 0 | 0 | 21 |
| Tom Gordon | LD | 1 | 0 | 0 | 0 | 0 | 0 | 0 | 0 | 21 |
| Daniel Francis | Lab | 8 | 1 | 0 | 0 | 0 | 0 | 1 | 0 | 22 |
| Danny Kruger | Con | 7 | 1 | 0 | 0 | 0 | 0 | 0 | 0 | 22 |
| Jack Abbott | Lab | 4 | 1 | 0 | 0 | 0 | 0 | 0 | 1 | 22 |
| Jack Abbott | Lab | 1 | 0 | 0 | 0 | 0 | 0 | 0 | 0 | 22 |
| Jake Richards |  | 4 | 0 | 0 | 0 | 0 | 0 | 0 | 0 | 22 |
| Kim Leadbeater | Lab | 7 | 1 | 0 | 0 | 0 | 0 | 0 | 0 | 22 |
| Kit Malthouse | Con | 4 | 1 | 0 | 0 | 0 | 0 | 0 | 0 | 22 |
| Lewis Atkinson | Lab | 3 | 0 | 0 | 0 | 0 | 0 | 0 | 1 | 22 |
| Naz Shah | Lab | 4 | 1 | 0 | 1 | 1 | 0 | 1 | 1 | 22 |
| Naz Shah | Lab | 1 | 0 | 0 | 0 | 0 | 0 | 0 | 0 | 22 |
| Rachel Hopkins | Lab | 1 | 0 | 0 | 0 | 0 | 0 | 0 | 0 | 22 |
| Rebecca Paul | Con | 18 | 1 | 1 | 1 | 1 | 0 | 1 | 1 | 22 |
| Sarah Olney | LD | 1 | 0 | 0 | 0 | 0 | 0 | 0 | 0 | 22 |
| The Chair |  | 3 | 0 | 0 | 0 | 0 | 0 | 0 | 0 | 22 |
| Tom Gordon | LD | 1 | 1 | 0 | 0 | 0 | 0 | 0 | 0 | 22 |
|  |  | 8 | 0 | 0 | 0 | 0 | 0 | 0 | 0 | 22 |
| Daniel Francis | Lab | 1 | 0 | 0 | 0 | 0 | 0 | 0 | 0 | 23 |
| Daniel Francis | Lab | 1 | 0 | 0 | 0 | 0 | 0 | 0 | 0 | 23 |
| Danny Kruger | Con | 47 | 1 | 0 | 1 | 1 | 0 | 1 | 1 | 23 |
| Marie Tidball | Lab | 1 | 0 | 0 | 0 | 0 | 0 | 0 | 0 | 23 |
| Opher |  | 1 | 0 | 0 | 0 | 0 | 0 | 0 | 0 | 23 |
| Simon Opher |  | 1 | 0 | 0 | 0 | 0 | 0 | 0 | 0 | 23 |
| Jack Abbott | Lab | 6 | 0 | 0 | 0 | 0 | 0 | 0 | 0 | 23 |
| Jack Abbott | Lab | 1 | 0 | 0 | 0 | 0 | 0 | 0 | 0 | 23 |
| Kim Leadbeater | Lab | 27 | 0 | 0 | 0 | 0 | 0 | 0 | 0 | 23 |
| Kit Malthouse | Con | 15 | 0 | 0 | 0 | 0 | 0 | 0 | 0 | 23 |
| Lewis Atkinson | Lab | 7 | 1 | 0 | 0 | 0 | 0 | 1 | 1 | 23 |
| Liz Saville Roberts | PC | 4 | 0 | 0 | 0 | 0 | 0 | 0 | 0 | 23 |
| Naz Shah | Lab | 17 | 0 | 1 | 0 | 1 | 1 | 0 | 1 | 23 |
| Naz Shah | Lab | 1 | 0 | 0 | 0 | 0 | 0 | 0 | 1 | 23 |
| Rebecca Paul | Con | 1 | 0 | 0 | 0 | 0 | 0 | 0 | 0 | 23 |
| Rebecca Paul | Con | 1 | 0 | 0 | 0 | 0 | 0 | 0 | 0 | 23 |
| Sarah Green | LD | 1 | 0 | 0 | 0 | 0 | 0 | 0 | 0 | 23 |
| Sarah Olney | LD | 8 | 0 | 0 | 0 | 0 | 0 | 1 | 0 | 23 |
| Sarah Sackman | | 28 | 0 | 0 | 0 | 1 | 0 | 0 | 0 | 23 |
| Sean Woodcock | Lab | 3 | 0 | 0 | 0 | 0 | 0 | 0 | 0 | 23 |
| Speaker name |  | 2 | 0 | 0 | 0 | 0 | 0 | 0 | 0 | 23 |
| Stephen Kinnock | Lab | 1 | 0 | 0 | 0 | 0 | 0 | 0 | 0 | 23 |
| The Chair |  | 10 | 0 | 0 | 0 | 0 | 0 | 0 | 0 | 23 |
| Tom Gordon | LD | 1 | 0 | 0 | 0 | 0 | 0 | 0 | 0 | 23 |
| Tom Gordon | LD | 1 | 0 | 0 | 0 | 0 | 0 | 0 | 0 | 23 |
|  |  | 2 | 0 | 0 | 0 | 0 | 0 | 0 | 0 | 23 |
| Daniel Francis | Lab | 2 | 0 | 0 | 0 | 0 | 0 | 0 | 0 | 24 |
| Daniel Francis | Lab | 1 | 0 | 0 | 0 | 0 | 0 | 0 | 0 | 24 |
| Danny Kruger | Con | 26 | 1 | 1 | 1 | 0 | 0 | 1 | 1 | 24 |
| Neil Shastri-Hurst | Con | 1 | 0 | 0 | 0 | 0 | 0 | 0 | 0 | 24 |
| Shastri-Hurst | Con | 4 | 0 | 0 | 0 | 0 | 0 | 0 | 0 | 24 |
| Kim Leadbeater | Lab | 22 | 0 | 0 | 0 | 0 | 0 | 1 | 1 | 24 |
| Kit Malthouse | Con | 6 | 1 | 0 | 0 | 0 | 0 | 0 | 0 | 24 |
| Liz Saville Roberts | PC | 1 | 0 | 0 | 0 | 0 | 0 | 0 | 0 | 24 |
| Naz Shah | Lab | 6 | 1 | 0 | 0 | 0 | 0 | 1 | 1 | 24 |
| Sarah Olney | LD | 1 | 0 | 0 | 0 | 0 | 0 | 0 | 0 | 24 |
| Sarah Olney | LD | 1 | 0 | 0 | 0 | 0 | 0 | 0 | 0 | 24 |
| Sean Woodcock | Lab | 1 | 0 | 0 | 0 | 0 | 0 | 0 | 0 | 24 |
| Sojan Joseph |  | 2 | 0 | 0 | 0 | 0 | 0 | 0 | 0 | 24 |
| Speaker name |  | 1 | 0 | 0 | 0 | 0 | 0 | 0 | 0 | 24 |
| Stephen Kinnock | Lab | 11 | 0 | 0 | 0 | 0 | 0 | 0 | 0 | 24 |
| The Chair |  | 9 | 0 | 0 | 0 | 0 | 0 | 0 | 0 | 24 |
| Daniel Francis | Lab | 2 | 0 | 0 | 0 | 0 | 0 | 0 | 0 | 25 |
| Danny Kruger | Con | 53 | 0 | 1 | 0 | 0 | 0 | 1 | 0 | 25 |
| Neil Shastri-Hurst | Con | 1 | 0 | 0 | 0 | 0 | 0 | 0 | 0 | 25 |
| Opher |  | 13 | 0 | 0 | 0 | 0 | 0 | 0 | 0 | 25 |
| Shastri-Hurst | Con | 5 | 0 | 0 | 0 | 0 | 0 | 0 | 0 | 25 |
| Simon Opher |  | 1 | 0 | 0 | 0 | 0 | 0 | 0 | 0 | 25 |
| Jack Abbott | Lab | 16 | 0 | 0 | 0 | 0 | 1 | 0 | 0 | 25 |
| Jack Abbott | Lab | 1 | 0 | 0 | 0 | 0 | 0 | 0 | 0 | 25 |
| Kim Leadbeater | Lab | 28 | 0 | 0 | 0 | 0 | 0 | 0 | 0 | 25 |
| Kit Malthouse | Con | 11 | 0 | 0 | 0 | 0 | 0 | 0 | 0 | 25 |
| Lewis Atkinson | Lab | 12 | 0 | 0 | 0 | 0 | 0 | 0 | 0 | 25 |
| Naz Shah | Lab | 18 | 0 | 0 | 0 | 0 | 0 | 0 | 0 | 25 |
| Naz Shah | Lab | 1 | 0 | 0 | 0 | 0 | 0 | 0 | 0 | 25 |
| Rachel Hopkins | Lab | 8 | 0 | 0 | 0 | 0 | 0 | 0 | 0 | 25 |
| Rachel Hopkins | Lab | 1 | 0 | 0 | 0 | 0 | 0 | 0 | 0 | 25 |
| Rebecca Paul | Con | 26 | 1 | 0 | 1 | 0 | 0 | 0 | 0 | 25 |
| Rebecca Paul | Con | 1 | 0 | 0 | 0 | 0 | 0 | 0 | 0 | 25 |
| Sarah Olney | LD | 10 | 0 | 0 | 0 | 0 | 0 | 0 | 0 | 25 |
| Sean Woodcock | Lab | 5 | 0 | 0 | 0 | 0 | 0 | 0 | 0 | 25 |
| Sean Woodcock | Lab | 1 | 0 | 0 | 0 | 0 | 0 | 0 | 0 | 25 |
| Sojan Joseph |  | 6 | 0 | 0 | 0 | 0 | 0 | 0 | 0 | 25 |
| Speaker name |  | 8 | 0 | 0 | 0 | 0 | 0 | 0 | 0 | 25 |
| Stephen Kinnock | Lab | 29 | 0 | 0 | 0 | 1 | 0 | 0 | 0 | 25 |
| The Chair |  | 24 | 0 | 0 | 0 | 0 | 0 | 0 | 0 | 25 |
| Tom Gordon | LD | 3 | 0 | 0 | 0 | 0 | 0 | 0 | 0 | 25 |
| Tom Gordon | LD | 1 | 0 | 0 | 0 | 0 | 0 | 0 | 0 | 25 |
|  |  | 18 | 0 | 0 | 0 | 0 | 0 | 0 | 0 | 25 |
| Danny Kruger | Con | 12 | 0 | 0 | 1 | 0 | 0 | 0 | 0 | 26 |
| Marie Tidball | Lab | 1 | 0 | 0 | 0 | 0 | 0 | 0 | 0 | 26 |
| Tidball | Lab | 3 | 0 | 0 | 1 | 0 | 0 | 0 | 0 | 26 |
| Jack Abbott | Lab | 3 | 0 | 0 | 1 | 0 | 0 | 0 | 0 | 26 |
| Jack Abbott | Lab | 1 | 0 | 0 | 0 | 0 | 0 | 0 | 0 | 26 |
| Kim Leadbeater | Lab | 6 | 0 | 0 | 0 | 0 | 0 | 0 | 0 | 26 |
| Kit Malthouse | Con | 6 | 0 | 0 | 0 | 0 | 0 | 0 | 0 | 26 |
| Lewis Atkinson | Lab | 9 | 0 | 0 | 0 | 0 | 0 | 0 | 0 | 26 |
| Liz Saville Roberts | PC | 2 | 0 | 0 | 0 | 0 | 0 | 0 | 0 | 26 |
| Naz Shah | Lab | 11 | 0 | 0 | 0 | 0 | 0 | 0 | 0 | 26 |
| Naz Shah | Lab | 1 | 0 | 0 | 0 | 0 | 0 | 0 | 0 | 26 |
| Rachel Hopkins | Lab | 1 | 0 | 0 | 0 | 0 | 0 | 0 | 0 | 26 |
| Rebecca Paul | Con | 3 | 0 | 0 | 0 | 0 | 0 | 0 | 0 | 26 |
| Rebecca Paul | Con | 1 | 0 | 0 | 0 | 0 | 0 | 0 | 0 | 26 |
| Speaker name |  | 1 | 0 | 0 | 0 | 0 | 0 | 0 | 0 | 26 |
| Stephen Kinnock | Lab | 2 | 0 | 0 | 0 | 0 | 0 | 0 | 0 | 26 |
| The Chair |  | 1 | 0 | 0 | 0 | 0 | 0 | 0 | 0 | 26 |
|  |  | 1 | 0 | 0 | 0 | 0 | 0 | 0 | 0 | 26 |
| Daniel Francis | Lab | 6 | 0 | 0 | 0 | 0 | 0 | 0 | 1 | 27 |
| Danny Kruger | Con | 40 | 0 | 0 | 0 | 0 | 0 | 0 | 0 | 27 |
| Marie Tidball | Lab | 1 | 0 | 0 | 0 | 0 | 0 | 0 | 0 | 27 |
| Tidball | Lab | 6 | 0 | 0 | 0 | 0 | 0 | 0 | 0 | 27 |
| Jack Abbott | Lab | 2 | 0 | 0 | 0 | 0 | 1 | 0 | 0 | 27 |
| Jack Abbott | Lab | 1 | 0 | 0 | 0 | 0 | 0 | 0 | 0 | 27 |
| Jake Richards |  | 4 | 0 | 0 | 0 | 0 | 0 | 0 | 0 | 27 |
| Kim Leadbeater | Lab | 33 | 1 | 0 | 0 | 0 | 0 | 1 | 0 | 27 |
| Kit Malthouse | Con | 7 | 0 | 0 | 0 | 0 | 0 | 0 | 0 | 27 |
| Lewis Atkinson | Lab | 6 | 0 | 0 | 0 | 0 | 0 | 0 | 0 | 27 |
| Naz Shah | Lab | 12 | 1 | 0 | 0 | 0 | 0 | 0 | 0 | 27 |
| Naz Shah | Lab | 1 | 1 | 0 | 0 | 0 | 0 | 0 | 0 | 27 |
| Rebecca Paul | Con | 7 | 1 | 0 | 0 | 1 | 1 | 0 | 0 | 27 |
| Rebecca Paul | Con | 1 | 0 | 0 | 0 | 0 | 0 | 0 | 0 | 27 |
| Sarah Olney | LD | 5 | 0 | 0 | 0 | 0 | 1 | 0 | 0 | 27 |
| Sarah Olney | LD | 1 | 0 | 0 | 0 | 0 | 0 | 0 | 0 | 27 |
| Sarah Sackman | | 15 | 1 | 0 | 0 | 0 | 0 | 0 | 0 | 27 |
| Sean Woodcock | Lab | 6 | 0 | 0 | 0 | 0 | 0 | 0 | 0 | 27 |
| Sean Woodcock | Lab | 1 | 0 | 0 | 0 | 0 | 0 | 0 | 0 | 27 |
| Sojan Joseph |  | 1 | 0 | 0 | 0 | 0 | 0 | 0 | 0 | 27 |
| Speaker name |  | 6 | 0 | 0 | 0 | 0 | 0 | 0 | 0 | 27 |
| Stephen Kinnock | Lab | 10 | 0 | 0 | 0 | 0 | 0 | 0 | 0 | 27 |
| The Chair |  | 34 | 0 | 0 | 0 | 0 | 0 | 0 | 0 | 27 |
| Tom Gordon | LD | 1 | 0 | 0 | 0 | 0 | 0 | 0 | 0 | 27 |
|  |  | 5 | 0 | 0 | 0 | 0 | 0 | 0 | 0 | 27 |
| Daniel Francis | Lab | 1 | 0 | 0 | 0 | 0 | 0 | 0 | 0 | 28 |
| Danny Kruger | Con | 10 | 0 | 1 | 0 | 1 | 0 | 0 | 0 | 28 |
| Danny Kruger | Con | 1 | 0 | 0 | 0 | 0 | 0 | 0 | 0 | 28 |
| Opher |  | 6 | 0 | 0 | 0 | 1 | 0 | 0 | 0 | 28 |
| Simon Opher |  | 1 | 0 | 0 | 0 | 0 | 0 | 0 | 0 | 28 |
| Kim Leadbeater | Lab | 6 | 0 | 0 | 0 | 0 | 0 | 0 | 0 | 28 |
| Kit Malthouse | Con | 4 | 0 | 0 | 0 | 0 | 0 | 0 | 0 | 28 |
| Lewis Atkinson | Lab | 7 | 0 | 0 | 0 | 0 | 0 | 0 | 0 | 28 |
| Liz Saville Roberts | PC | 3 | 0 | 0 | 0 | 0 | 0 | 0 | 0 | 28 |
| Naz Shah | Lab | 4 | 0 | 0 | 0 | 0 | 0 | 0 | 0 | 28 |
| Naz Shah | Lab | 1 | 1 | 0 | 0 | 1 | 1 | 1 | 0 | 28 |
| Rachel Hopkins | Lab | 1 | 0 | 0 | 0 | 0 | 0 | 0 | 0 | 28 |
| Rebecca Paul | Con | 5 | 0 | 0 | 1 | 0 | 0 | 0 | 0 | 28 |
| Sarah Olney | LD | 17 | 0 | 1 | 1 | 1 | 0 | 0 | 1 | 28 |
| Sean Woodcock | Lab | 3 | 0 | 0 | 0 | 0 | 0 | 0 | 0 | 28 |
| Speaker name |  | 1 | 0 | 0 | 0 | 0 | 0 | 0 | 0 | 28 |
| The Chair |  | 3 | 0 | 0 | 0 | 0 | 0 | 0 | 0 | 28 |
| Tom Gordon | LD | 1 | 0 | 0 | 0 | 0 | 0 | 0 | 0 | 28 |
|  |  | 1 | 0 | 0 | 0 | 0 | 0 | 0 | 0 | 28 |
| Daniel Francis | Lab | 12 | 0 | 0 | 0 | 0 | 0 | 0 | 1 | 29 |
| Danny Kruger | Con | 26 | 1 | 0 | 0 | 1 | 0 | 0 | 0 | 29 |
| Marie Tidball | Lab | 1 | 0 | 0 | 0 | 1 | 0 | 0 | 0 | 29 |
| Neil Shastri-Hurst | Con | 1 | 0 | 0 | 0 | 0 | 0 | 0 | 0 | 29 |
| Opher |  | 4 | 0 | 0 | 0 | 0 | 0 | 0 | 0 | 29 |
| Simon Opher |  | 1 | 0 | 0 | 0 | 0 | 0 | 0 | 0 | 29 |
| Tidball | Lab | 10 | 1 | 0 | 0 | 1 | 0 | 0 | 0 | 29 |
| Jack Abbott | Lab | 4 | 0 | 0 | 0 | 0 | 0 | 0 | 0 | 29 |
| Jack Abbott | Lab | 1 | 1 | 0 | 0 | 0 | 0 | 0 | 0 | 29 |
| Jake Richards |  | 3 | 0 | 0 | 0 | 0 | 0 | 0 | 0 | 29 |
| Kim Leadbeater | Lab | 47 | 0 | 0 | 0 | 0 | 0 | 0 | 0 | 29 |
| Kit Malthouse | Con | 6 | 0 | 0 | 0 | 0 | 0 | 0 | 0 | 29 |
| Lewis Atkinson | Lab | 6 | 0 | 0 | 0 | 0 | 0 | 0 | 0 | 29 |
| Liz Saville Roberts | PC | 6 | 0 | 0 | 0 | 0 | 0 | 0 | 0 | 29 |
| Naz Shah | Lab | 25 | 1 | 0 | 1 | 1 | 1 | 0 | 0 | 29 |
| Naz Shah | Lab | 1 | 0 | 0 | 0 | 0 | 0 | 0 | 0 | 29 |
| Rachel Hopkins | Lab | 1 | 0 | 0 | 0 | 0 | 0 | 0 | 0 | 29 |
| Rebecca Paul | Con | 16 | 1 | 0 | 0 | 0 | 0 | 1 | 1 | 29 |
| Rebecca Paul | Con | 1 | 0 | 0 | 0 | 0 | 0 | 0 | 0 | 29 |
| Sarah Olney | LD | 13 | 0 | 0 | 0 | 1 | 1 | 0 | 1 | 29 |
| Sarah Sackman | | 7 | 1 | 1 | 0 | 0 | 0 | 0 | 0 | 29 |
| Speaker name |  | 11 | 0 | 0 | 0 | 0 | 0 | 0 | 0 | 29 |
| Stephen Kinnock | Lab | 29 | 0 | 0 | 0 | 0 | 0 | 0 | 0 | 29 |
| The Chair |  | 28 | 1 | 0 | 0 | 0 | 0 | 0 | 0 | 29 |
| Tom Gordon | LD | 1 | 0 | 0 | 0 | 0 | 0 | 0 | 0 | 29 |

### Supplementary file S.4. Correlations matrices

| Reading |  |  |  |  |  |  |  |
| --- | --- | --- | --- | --- | --- | --- | --- |
|  | Family | Medical | Poor care | Economic | Ethnic | Self | Mental |
| Family | 1.00 | 0.76 | 0.40 | 0.57 | 0.26 | 0.50 | 0.54 |
| Medical | 0.76 | 1.00 | 0.53 | 0.46 | 0.08 | 0.46 | 0.60 |
| Poor care | 0.40 | 0.53 | 1.00 | 0.50 | 0.31 | 0.39 | 0.56 |
| Economic | 0.57 | 0.46 | 0.50 | 1.00 | 0.38 | 0.52 | 0.57 |
| Ethnic | 0.26 | 0.08 | 0.31 | 0.38 | 1.00 | 0.10 | 0.31 |
| Self | 0.50 | 0.46 | 0.39 | 0.52 | 0.10 | 1.00 | 0.43 |
| Mental | 0.54 | 0.60 | 0.56 | 0.57 | 0.31 | 0.43 | 1.00 |
| Sitting |  |  |  |  |  |  |  |
|  | Family | Medical | Poor care | Economic | Ethnic | Self | Mental |
| Family | 1.00 | 0.17 | 0.23 | 0.36 | 0.15 | 0.45 | 0.25 |
| Medical | 0.17 | 1.00 | 0.31 | 0.16 | 0.21 | 0.21 | 0.49 |
| Poor care | 0.23 | 0.31 | 1.00 | 0.36 | 0.26 | 0.28 | 0.26 |
| Economic | 0.36 | 0.16 | 0.36 | 1.00 | 0.30 | 0.35 | 0.20 |
| Ethnic | 0.15 | 0.21 | 0.26 | 0.30 | 1.00 | 0.11 | 0.20 |
| Self | 0.45 | 0.21 | 0.28 | 0.35 | 0.11 | 1.00 | 0.33 |
| Mental | 0.25 | 0.49 | 0.26 | 0.20 | 0.20 | 0.33 | 1.00 |
